# Efficacy-adjusted use: modelling a refined metric of insecticide treated net coverage across Africa

**DOI:** 10.64898/2026.08.13.26360347

**Authors:** Eugene Tan, Rubi Jayaseelen, Adam Saddler, Mauricio van den Berg, Camilo Vargas, Nick Golding, Daniel J. Weiss, Amelia Bertozzi-Villa, Peter W. Gething, Tasmin L. Symons

## Abstract

Insecticide-treated net (ITN) use – defined as the proportion of a population that use ITNs – is a measure of ITN uptake that is used in the estimation of malaria burden and evaluation of intervention programs. However, binary classification of individuals as users or non-users does not account for variations in ITN-related protection attributed to deleterious factors such as chemical and physical degradation, and increased insecticide resistance in vector populations. In this paper, we present a parsimonious model for malaria dynamics in mosquito-human populations in the presence of varying ITN use conditions. Using this model, we propose a new standardised measure of ITN coverage termed the “efficacy-adjusted use” defined as the equivalent level of use, assuming fully efficacious nets, that would be required to achieve the same level of theoretical EIR reduction. This more nuanced measure is used as a proxy for studying ITN-attributed protection across 44 African countries. We find that estimated protection levels in current ITN paradigms is significantly lower than indicated by crude ITN use metrics, with insecticide resistance having the largest deleterious effect. Furthermore, recent adoption of next-generation nets is estimated to have mitigated a 13% reduction in protection compared to a counterfactual pyrethroid only scenario.

## I. INTRODUCTION

While malaria remains a leading cause of illness and death in sub-Saharan Africa, the current disease landscape reflects significant progress over the past 25 years, with rates of malaria incidence and mortality decreasing by around 20% and 50% respectively over that time [1, 2]. Much of this progress in reducing malaria burden is attributed to the implementation of various intervention measures such as indoor residual spraying (IRS), artemisinin combination therapy (ACT), and insecticide treated nets (ITN) [3].

Among the above three main classes of interventions, ITNs have been the most impactful form of intervention against malaria infection and clinical incidence and account for an estimated 68% of averted cases from 2000-2015 [3]. The widespread adoption of ITNs starting from the early-mid 2000s coincided with large decreases in malaria incidence and parasite rates. Starting at a case incidence of 79 cases per 1000 among countries at risk in 2000, concerted efforts to combat malaria resulted in a decrease in incidence to 68 cases per 1000 by 2010. This progress continued through the mid 2010s with incidence reaching as low as 59 cases per 1000 among countries at risk. However, recent years have seen a reversal in this trend with current estimates indicating a slow but steady uptick in case incidence to 64 cases per 1000 [1]. The concerning trends in malaria incidence coincide with the stagnation in ITN coverage, which has remained at a continental average of around 50% popuation-level use since 2015 [4] and continues to falls short of the WHO recommended level of 80% coverage [5, 6]. These trends are further worsened by the increasing prevalence of insecticide resistance (IR) among malaria vector populations [7–10]. This has led to the development and adoption of more durable and bioefficacious next-generation LLINs. Examples include pyrethroid-piperonyl butoxide (PBO) and dual active ingredient (DAI) net types, whose total market share of net crop has grown to 34% in 2024 [4].

Increasing biological and economical constraints on malaria interventions coupled with a rapidly changing epidemiological landscape has increased the demand and highlighted the importance of ITN coverage models as a component in data-driven planing for optimal impact[4, 11–13]. The outputs of these models include coverage metrics such as nets-per-capita (NPC), access and use. These measures track the progress of ITN adoption, and inform the planning and optimisation of ITN distributions by enabling the simulation of various campaign strategies to identify and address regions of highest need. They are also employed in downstream modelling efforts to better understand the impact of ITN interventions. Coverage metrics are critical for guiding various stages of the subnational tailoring process for malaria programs [14, 15]. ITN coverage models also provide information on the protection level of communities and are critical components for modelling and estimating malaria infection prevalence and disease burden [16, 17].

The current state-of-the-art spatial model for ITN coverage is the Multitype-ITN model (MITN) proposed by *Tan et al* [4, 13]. This model combines a compartmental stock-and-flow model with geospatial disaggregation to estimate net crop composition by age and type - referred to as net demography - and tracks standard coverage metrics (NPC, access, use) across space and time. One of the most important coverage metrics is ITN use, which is defined as the proportion of a population who are active net users. Estimates of ITN use provide a descriptive proxy for the potential impact of ITNs on malaria transmission and are used in various downstream analyses for estimating parasite rates and disease burden [16]. Whilst the MITN model tracks net demography, crude estimates of ITN use are a dichotomised measure of individuals’ utilisation of nets and do not account for factors that may affect the efficacy of a net being used. Two such deleterious effects are age-related waning bioefficacy, and reduced protection due to insecticide resistance among local vector populations. As a result, crude ITN use does not capture important variation in ITN effectiveness, resulting in unresolved model errors in downstream applications between regions or times with similar use, but differing ITN age and and local vector characteristics.

In this paper, we present a method for estimating a more nuanced measure of ITN use that accounts for the effects of net age and insecticide resistance. We combine existing models of insecticide susceptibility, and ITN durability with a modified delay differential equation (DDE) compartmental model to provide a parsimonious model for calculating a theoretical notion of entomological inoculation rate (EIR). Theoretical values of EIR are used as surrogates to calculate penalty factors for ITN use. These penalties are used to calculate an efficacy-adjusted use – the level of use of fully efficacious brand new nets that will provide an equivalent level of EIR reduction – which accounts for age-related degradation in bioefficacy and IR effects. In our analyses, we use efficacy-adjusted use as a proxy for the level of protection against infectious bites in a population. The proposed method is used to calculate efficacy-adjusted use across 44 countries in Africa, thus providing several key epidemiological insights on real ITN-related protection and the impact of net age and IR over time.

## II. INSECTICIDE TREATED NETS (ITNS)

Insecticide-treated nets (ITNs) consist of a synthetic or cotton woven net whose fibres are impregnated with insecticidal agents. As one of the most widely used malaria interventions, they provide protection via two main mechanisms. Firstly, ITNs directly reduce the frequency of mosquito-human contact via physical barrier and chemical deterrence effects, which results in decreased infection risk for net-users. Secondly, insecticides reduce vector populations by killing any mosquitoes that come in contact with the net. In addition to providing direct protection to users, ITNs – when used at a sufficiently high coverage – also confer community level protection to nearby non-users as well [18–21]. We note that apart from these two main mechanisms, some special cases of dual active ingredient nets work to reduce fecundity instead of directly killing mosquito vectors, such as those where pyriproxiphen is used. However, for model simplcity we do not model this case in this paper.

Excluding the non-insecticidal cotton nets used prior to 2000, which provided only a barrier effect or protection, ITNs may be broadly separated into three main classes. (1) Conventional ITNs (cITNs) are non-washable insecticide coated nets that were widely used prior to 2010. (2) Long-lasting Insecticide Nets (LLINs) are a more durable and washable insecticide nets, which gained rapid adoption from 2010 and remain the predominant type of ITNs accounting for two-thirds of Africa’s usage market share as of 2024. (3) Next-Generation LLINs are a recently developed class of insecticidal nets that have been increasingly adopted in response to growing insecticide resistance, and has experience steadily growing market share since 2019 [4]. Two of the most common types are pyrethroid-piperonyl butoxide (PBO) nets, and dual active ingredient (DAI). PBO nets partially restore insecticide susceptibility by combining traditional pyrethroid insecticides with piperonyl butoxide, which suppresses enzymes in mosquitoes that inhibit the effect of pyrethroids [15, 22]. In contrast, DAI nets combine two insecticide classes that exploit different biological pathways to overcome insecticide resistance [23].

## III. ESTIMATES OF ITN USE

ITN use is defined as the proportion of people in a given population that use an ITN and previous estimates are often agnostic of net age and type. Estimates of ITN use and other coverage metrics can be estimated at specfic points in time from a combination of household surveys of net ownership, manufacturer delivery data and reported distribution data [4, 11]. However, the sampling frequency and sparsity of available data across various countries do not allow direct estimates to be easily calculated [4]. Instead, estimates of ITN coverage have used a mathematical model to corroborate multiple datasets and interpolate between observed data points [24, 25].

The Multitype-ITN (MITN) model proposed by *Tan et al*. is one such model that is employed to produce ITN coverage estimates. The MITN model reconciles various data sources consisting of geolocated household surveys of net ownership and household demography, net distribution, net delivery and population data to construct estimates of ITN coverage metrics (NPC, access, use). These metrics are uniformly resolved across space and time. In this section, we provide a brief overview of the MITN model for calculating estimates of ITN use from survey and observational data. For more detail, we refer to the original work by *Tan et al*.

The MITN model consists of three components that are run sequentially to produce spatiotemporal estimates of ITN use. Mathematically speaking, the model attempts to calculate the variation of a coverage metric over some spatial and temporal domain. These three components are the stock-and-flow model, NPC-Access conversion model, and spatiotemporal disaggregation model (see Figure 1).

**FIG. 1.**
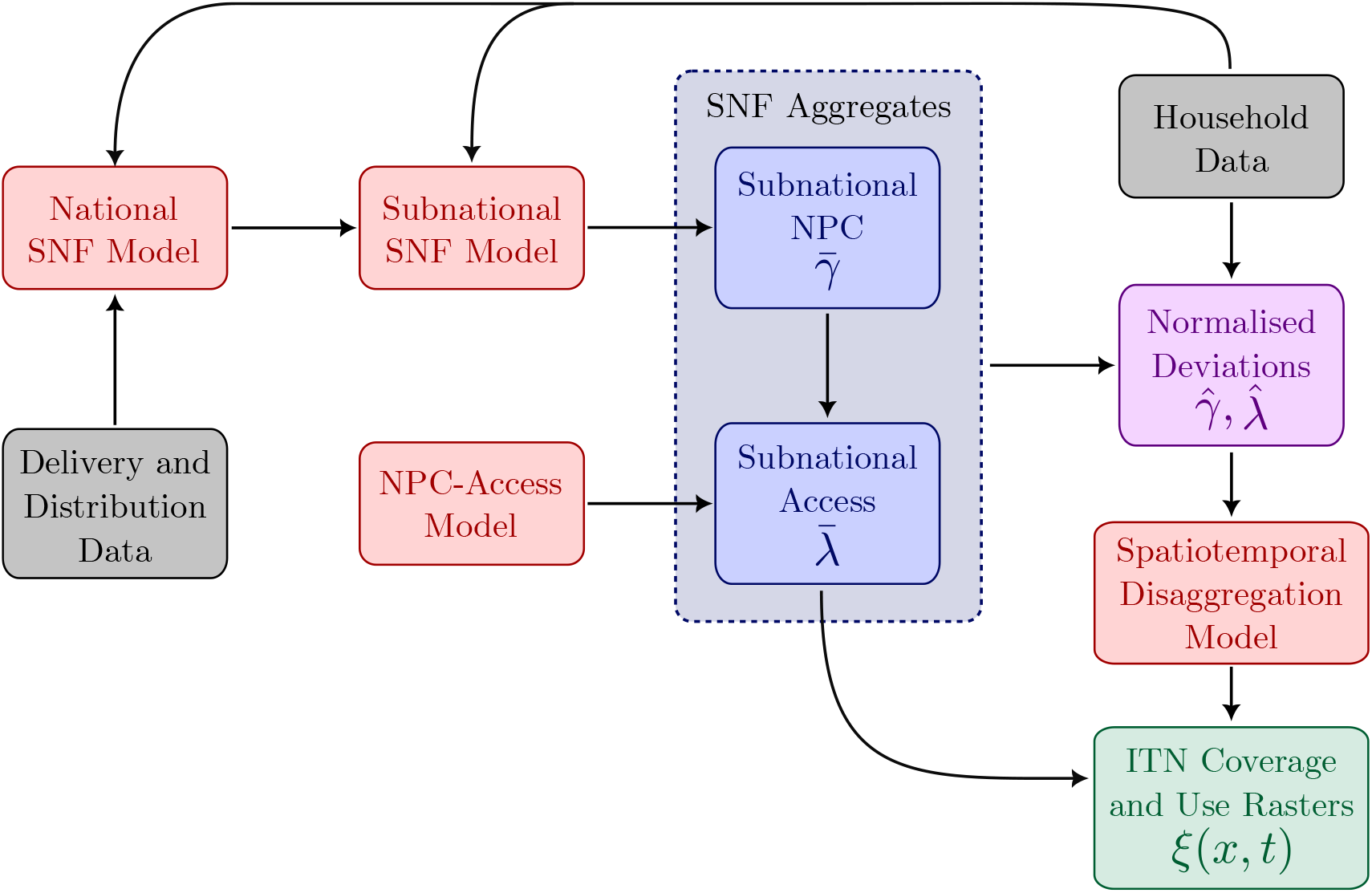
Schematic of the MITN model by *Tan et al*. for estimating ITN coverage. Box colours correspond to input data (grey), model components (red), intermediate variables (magenta) and ITN coverage outputs (green and blue).

The stock-and-flow (SNF) model is a compartmental model that simulates the stockpile, distribution and attrition in net volumes within individual countries separated according net type (cITN, LLIN, PBO, DAI). The SNF model is calibrated against a collection of household surveys, national distribution and manufacturer delivery data to provide posterior estimates of net attrition parameters. These parameters are used to describe the rates at which ITNs enter and disappear from the net crop of a community.

Once calibrated, learned attrition parameters for each net type can be used to estimate the composition of a region’s net crop by type and age [4]. For net type, this breakdown is largely determined by the type and volume of nets that are distributed and is incorporated through the inclusion of WHO net distribution data. Estimates of net age are calculated based on the posterior fits of net attrition. However, a limitation of the MITN model is that it only accounts for public sector distributions and does not include household acquisition of nets through the private sector. This assumption is valid during time periods following a mass campaign, but may be increasingly inaccurate for periods where government or public distribution is low and households attempt to acquire nets via the private sector. Despite this, model estimates for aggregates of ITN mean net age show that on a continent level broadly agree with that of those reported by surveys (MITN model = 1.25 years, DHS surveys = 1.48 years). Considering the difficulty and uncertainty for households to recall the age of individual nets they possess, the broad agreement between model and survey estimates provide confidence in the latter being a relatively reliable initial estimate of net age for use in our analyses.

Net access is defined as the proportion of people with adequate access to a net, where access is defined as a maximum of two people per net. The NPC-Access conversion model is used to convert estimates of nets-per-capita (NPC) into net access. This is done by learning the joint distribution of household size and nets per household using household survey data.

The SNF and NPC-Access conversion models only provide regional aggregates of ITN ownership and access. In practice, ITN coverage is spatially heterogeneous and can vary greatly even within the same national boundary. To capture this spatial variation, a spatiotemporal disaggregation model is used to disaggregate previously calculated values of NPC and access into a high resolution spatiotemporal grid. Grids for NPC and access are combined with geolocated survey observations of household use to produce spatiotemporal estimates of ITN use *ξ*(*x, t*). Final aggregate values for NPC, access and use can then be calculated using population weighted spatial averages.

## IV. COMPONENT MODELS AND DATA

### A. Insecticide resistance

The rise of insecticide resistance, and the associated increase in vector survival upon exposure to ITNs, has threatened the effectiveness of ITN campaigns. Previously, geospatial models of vector susceptibility were used to map insecticide resistance (pyrethroids and DDT) across multiple regions and years [7]. This approach was particularly effective in producing high resolution spatial estimates of insecticide resistance across domains (space and time) with reasonable amounts of data. However, this poses a challenge for calculating penalties for ITN use, for which data and model estimates spans across a larger domain. To represent vector susceptibility to insecticides, we employ a Bayesian hierarchical semi-mechanistic model proposed by *Golding et al*. We provide a brief summary of this model and refer readers to [26] for details.

Briefly, *Golding et al*. combines a geospatial statistics approach with mechanistic processes to model resistance growth and trait evolution for the purposes of inferring vector susceptibility to insecticides. This model of insecticide resistance consists of three submodel components: the observation model, trait evolution, and selection pressures.

The observation model is used to describe bioassay data of mosquito mortality collected in various experimental trials used to track insecticide resistance in a given region. A difference equation with a driving term is used to represent the evolution of traits conferring resistance due to various dynamic selection pressures. This equation is solved for each modelled 5×5 km pixel location with respect to statistically inferred values for selection pressures. A linear model with spatiotemporal covariates is used to represent spatiotemporal variations in selection pressures *w*. A collection of spatiotemporal covariates are used in the regression and fall within three main classes: vector control, agricultural crops, and human population level. Selection effects are modelled with a doubly hierarchical model with penalised complexity priors. The model is fitted against bioassay data from the IR Mapper [27, 28] and WHO Malaria Threats Map [29]. Outputs of the model comes in the form of a spatiotemporal scalar field of susceptibility given by *γ*(*x, t*) at location *x* and time *t*, which combines individually modelled insecticides using in LLINs and is defined at a 5×5km spatial resolution and an annual temporal resolution.

### B. Killing and deterrence effects

Active chemical agents in ITNs can affect mosquito behaviours via two modelled mechanisms: killing of mosquitoes due to physical contact with insecticide, and an indirect deterrence effect which reduces the frequency of nighttime bites [30, 31]. To account for this dual mode of action in our model, we define two parameters to describe the killing and deterrence effects afforded by the use of ITNs. The killing probability (*η*_*k*_) is the probability than a mosquito dies due to an attempted bite on an ITN user. Similarly, the deterrence ratio (*η*_*d*_) is the proportion of bites that are not averted due to the presence of an ITN (i.e. 1 *− η*_*d*_ is equivalent to the reduction in the number of bites).

We model killing and deterrence parameters (*η*_*k*_, *η*_*d*_) as a function of waning insecticide bioefficacy due to net age, and reduced vector susceptibility from insecticide resistance. To do so, we construct a simple Weibull relationship to represent the natural decay of bioefficacy over time,

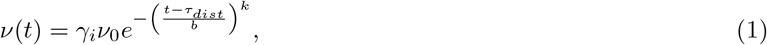

where *t − τ*_*dist*_ is the time since net distribution, *v*(*t*) is the insecticide bioefficacy, *v*_0_ = 1 is the baseline bioassay efficacy of a brand new net against a fully susceptible vector, and *γ*_*i*_ is the vector susceptibility of the given net type *i* as defined in Section IV A. Model parameters *b* and *k* are fitted against data compiled from a collection of longitudinal studies ITN 24 hour bioassay mortality from 2016-2024 [32–35]. For both parameters, we use a wide prior Gamma *∼* (9*/*2, 3*/*2) and calculate a MAP estimate of *b* = 2.45 and *k* = 3.8, with a bioefficacy of half-life of 2.22 years. Furthermore, for simplification we assume that all net types exhibit the same natural decay in bioefficacy over time in the absence of insecticide resistance.

While 24 hour bioassay mortality rates are an indicator of insecticide bioefficacy, they are evaluated in idealised conditions and are not equal to the performance of ITNs when deployed in the field represented, which we represent by *η*_*k*_ and *η*_*d*_. Results from experimental hut trial (EHT) studies across sub-Saharan Africa have indicated in-field reductions in bite rates that are lower than suggested by bioassay mortality. To convert between bioassay mortality and in-field estimates of killing and deterrence effects, we employ a two-part statistical model proposed by *Nash et al. [31],*

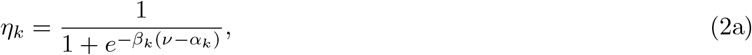

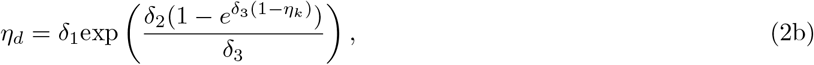

where *β*_*k*_, *α*_*k*_, *δ*_1_, *δ*_2_, *δ*_3_ are fitted parameters calibrated against EHT data (see Supplementary Information).

Because killing and deterrence effects are given as a function of net age and type, variations in the type and age composition of the deployed net crop must be accounted for. For this, we utilise the joint age and type net demography distributions output by the MITN model to calculate weighted average values of deterrence and killing parameters. This is given by

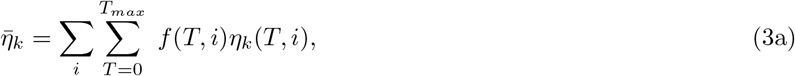

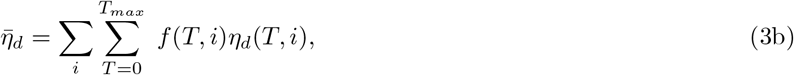

where *f* (*T, i*) is the fraction of nets across that are of age *T* and type *i*, and *η*_*k*_(*T, i*) and *η*_*d*_(*T, i*) are the killing and deterrence effects respectively.

### C. Barrier effects

ITNs can also contribute to the protection of an individual from bites by acting as a physical barrier. Prior to the widespread usage of ITNs, untreated nets were commonly used to provide some degree of protection against entomological inoculation [21]. Similarly, the usage of ITNs whose bioefficacy has been compromised should still afford some degree of protection based on the physical condition of the net.

Several studies have been conducted to model the physical degradation of ITNs over time. These measures track various factors such as hole size [36], abrasion damage and physical strength [37]. Models for ITN barrier effects should ideally account for these physical factors. Since successful entomological inoculation in the presence of a barrier is predominantly stochastic process, we argue that models with added complexity to account for physical degradation mechanisms (e.g. hole size) may not be easily tractable. To achieve a balance between model complexity and flexibility, we instead include a constant parameter *η*_*b*_ - termed the blocking probability - which is used to penalize the bite rate *α* experienced by an ITN user. This value is calibrated against models by *Unwin et al*. that attempt to disentangle barrier and insecticidal effects on EIR reduction. Details on this process is provided in the Supplementary Information [21].

## V. EIR SURROGATE MODELS

Our proposed method for calculating efficacy-adjusted estimates of ITN use centres on surrogate models of entomological inoculation rate (EIR) – defined as the number of infectious bites received by an individual per unit time. Estimate the reduction in EIR for a population with varying levels of ITN usage regimes provides a useful proxy for the level of protection against infection afforded by said interventions. However, simulating EIR requires a dynamical model for simulation.

Multiple models exist for simulating EIR with well-established examples include the Imperial model (malariasimulation) by *Griffin et al*. [38, 39], EMOD by the Institute for Disease Modelling (IDM) [40], and OpenMalaria [41, 42]. All of these models are highly descriptive and incorporate a large number of biological mechanisms including mosquito life-cycle mechanics, human host infection dynamics and malaria interventions such as IRS and ITN use, to produce a large array of epidemiological features and outputs. One of these outputs is a simulated value of EIR, which has also been successfully utilised in other studies on ITN effects [21]. Whilst these outputs would be sufficient for our analyses, there are two aspects that warrant consideration.

**TABLE I.** Definition and chosen values for theoretical EIR model parameters with and without ITN interventions.

| Parameter | Description | Value |
| --- | --- | --- |
| $\alpha_0$ | Baseline average human bite rate (HBR) per mosquito, per person, per day [44] | 0.33 |
| $N_{m,0}$ | Baseline mosquito density (mosquitoes per capita) at steady state | 29 |
| $p_m$ | Probability of mosquito getting infected after blood-feed [43] | 0.24 |
| $p_h$ | Probability of human getting infected after blood-feed [43] | 0.022 |
| $d_{m,NAT}$ | Natural death rate of healthy mosquitoes[43] | 0.0477 |
| $d_{m,INF}$ | Inflated death rate of infected mosquitoes due to likely older age when infectious | 0.17 |
| $\tau_m$ | Average latent period of malaria in mosquitoes in days [45] | 4 |
| $\tau_h$ | Average latent period of malaria in humans in days [46] | 14 |
| $\mu_h$ | Natural recovery rate of humans [43] | 0.0035 |
| $\eta_k$ | Probability of mosquito killing on attempted blood feed | [0, 1] |
| $\eta_d$ | Probability of mosquito deterrence due to ITN use | [0, 1] |
| $\eta_b$ | Reduction ratio in mosquito bite rate due to ITN blocking effect [21] | 0.372 |

Firstly, the model does not readily incorporate age and type characteristics of ITNs when simulating EIR rates of individuals. Adding further modifications to existing models – with already high model complexity and number of parameters – to include ITN age and type compositions would be unwieldy and pose challenges when performing sensitivity analyses. Secondly, many of these models are computationally expensive due the need to simulate multiple interacting individuals to sufficiently capture disease dynamics. Due to the algorithmic repetition of our analyses, performing repeated simulations of agents is computationally costly. Despite these drawbacks, we note that these individual-based models are purpose built to produce a rich set of outputs of which only one component – EIR – is required in our analyses.

We base our analyses on the Imperial model by *Griffin et al*. and propose a simplification that trades off dynamical complexity for parsimony, while being sufficiently descriptive to capture the impacts of ITN interventions for various age and type classes on EIR. Specifically, we construct a modified Ross-Macdonald compartmental model to describe variations in infected human and mosquito populations under various ITN intervention scenarios. Theoretical EIR values under a given ITN intervention scenario are compared against a control where no interventions are used. This is used to identify reduction ratio that broadly quantifies the level of protection afforded by ITN usage.

Our mechanistic model establishes two key findings. Firstly, despite including dynamical processes across different time scales, approximations for the steady state solutions closely track those of the simulated trajectory of the differential equations (see Supplementary Information). Secondly, these steady state solutions of the proposed can be numerically estimated with a high degree of computational efficiency. Because important time-varying parameters related to ITN coverage and efficacy (i.e. insecticide resistance, killing, deterrence and use) are unlikely to show large variations across these short time scales, these steady state solutions can be used to rapidly calculate surrogate estimates of the theoretical transmission and disease states associated with a given ITN coverage status.

For clarity, a list of definitions for the set of model parameters Θ =*{α*_0_, *p*_*m*_, *p*_*h*_, *d*_*m,NAT*_ , *d*_*m,INF*_ , *τ*_*m*_, *τ*_*h*_, *µ*_*h*_*}* is given in Table V. Full derivations for solutions are given in the Supplementary Information. Rather than doing an end-to-end calibration of the model, we utilise results from existing data-driven modelling studies to select values for model parameters. These include natural mosquito death rates [43], daily human bite rates (HBR) [44], human-mosquito transmission probabilities [43], and human recovery rates [43]. Whilst there is debate on the effect of senescence and infection on the mosquito mortality, there exist findings on potential differences in the extrinsic incubation period (EIP) due to factors such as age, infection and temperature. Therefore to account for these difference in our model, we select an inflated rate of death for infected mosquitoes given by *d*_*m*.*INF*_ due to older age given their opportunity to incubate an infection.

### A. EIR model without intervention

We first model the dynamics of human and mosquito populations with malaria in the absence of interventions and constrained by the following assumptions:

- The number of human deaths due to malaria is small relative to total population 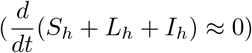
- Homogeneous mixing in all compartment groups

For this, we use a compartmental model with six compartments representing the susceptible, latent and infected populations of mosquitoes and humans respectively given by *S*_*m*_, *L*_*m*_, *I*_*m*_, *S*_*h*_, *L*_*h*_, *I*_*h*_ (see Figure 2). The dynamics between each compartments are given by the following delay differential equation,

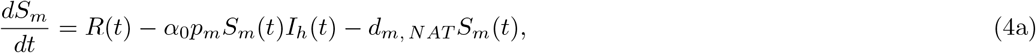

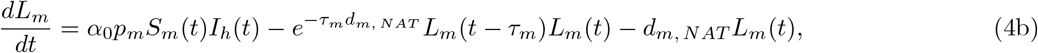

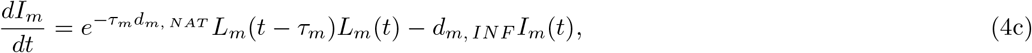

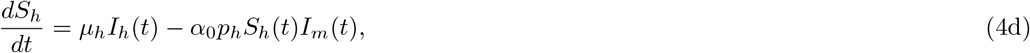

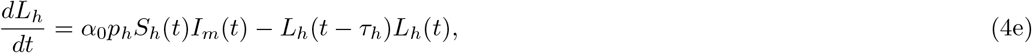

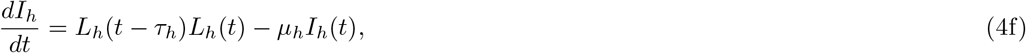

and we define the total populations of mosquitoes as *N*_*m*_ = *S*_*m*_ +*L*_*m*_ +*I*_*m*_ and human poulations given as a normalised proportion given as *N*_*h*_ = *S*_*h*_ + *L*_*h*_ + *I*_*h*_ = 1 respectively. For a given set of model parameters Θ, there exists a unique steady state 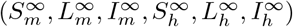 that can be numerically solved. This is used to calculate a baseline reproduction rate for mosquitoes *R*_*∞*_ = lim_*t → ∞*_ *R*(*t*) (see Supplementary Information). Similarly, the theoretical baseline EIR in the absence of interventions for a set of model parameters Θ can be calculated as

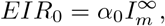

where *α*_0_ is the daily human bite rate (average number of daily bites per human per mosquito).

**FIG. 2.**
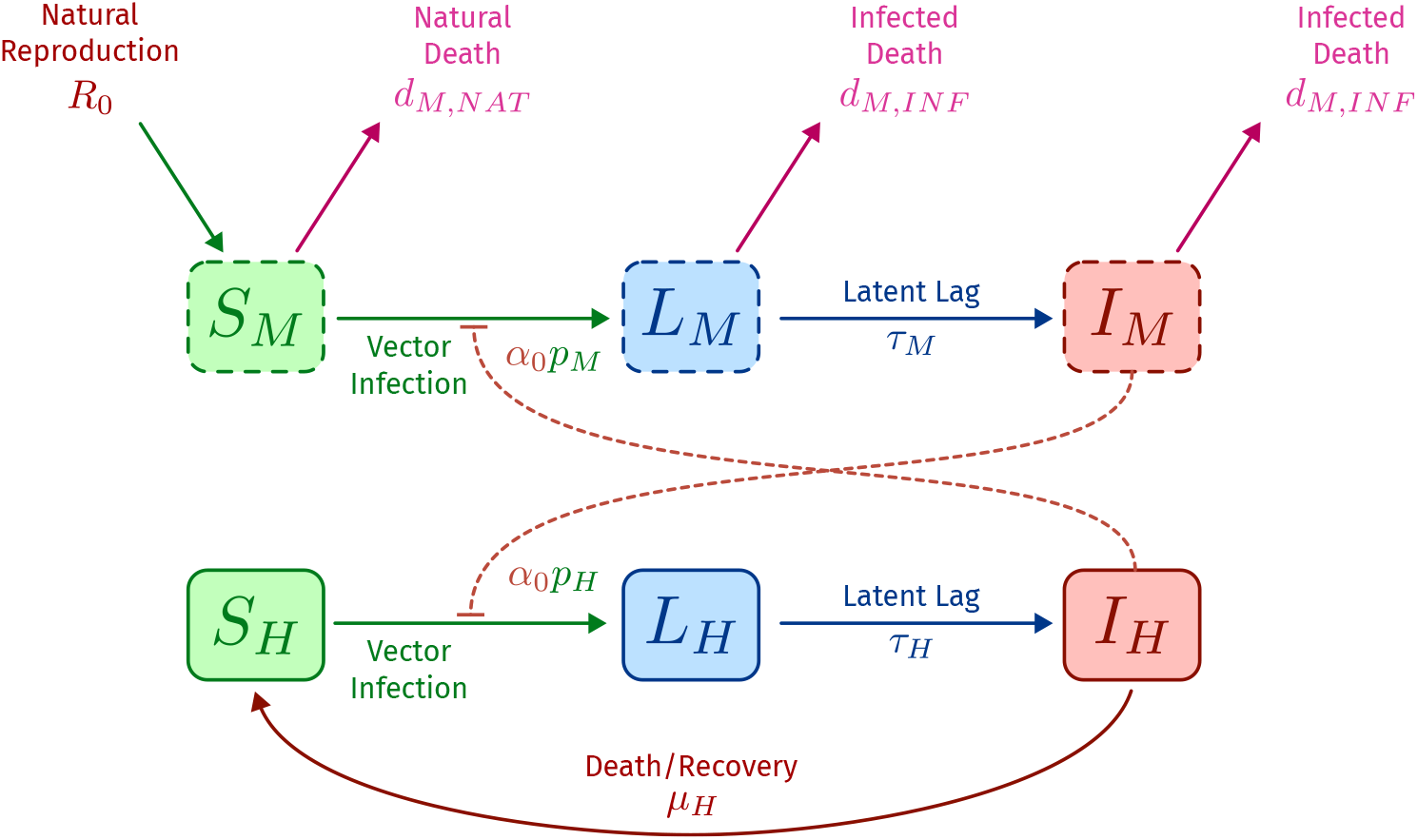
Schematic of the compartmental EIR model with delay in the absence of ITN interventions.

### B. EIR model with interventions

After calculating the baseline reproduction rate for mosquitoes in the control case, we apply it as a driving term for a modified compartmental model that includes intervention effects (killing, deterrence and barrier) (see Figure 3).

**FIG. 3.**
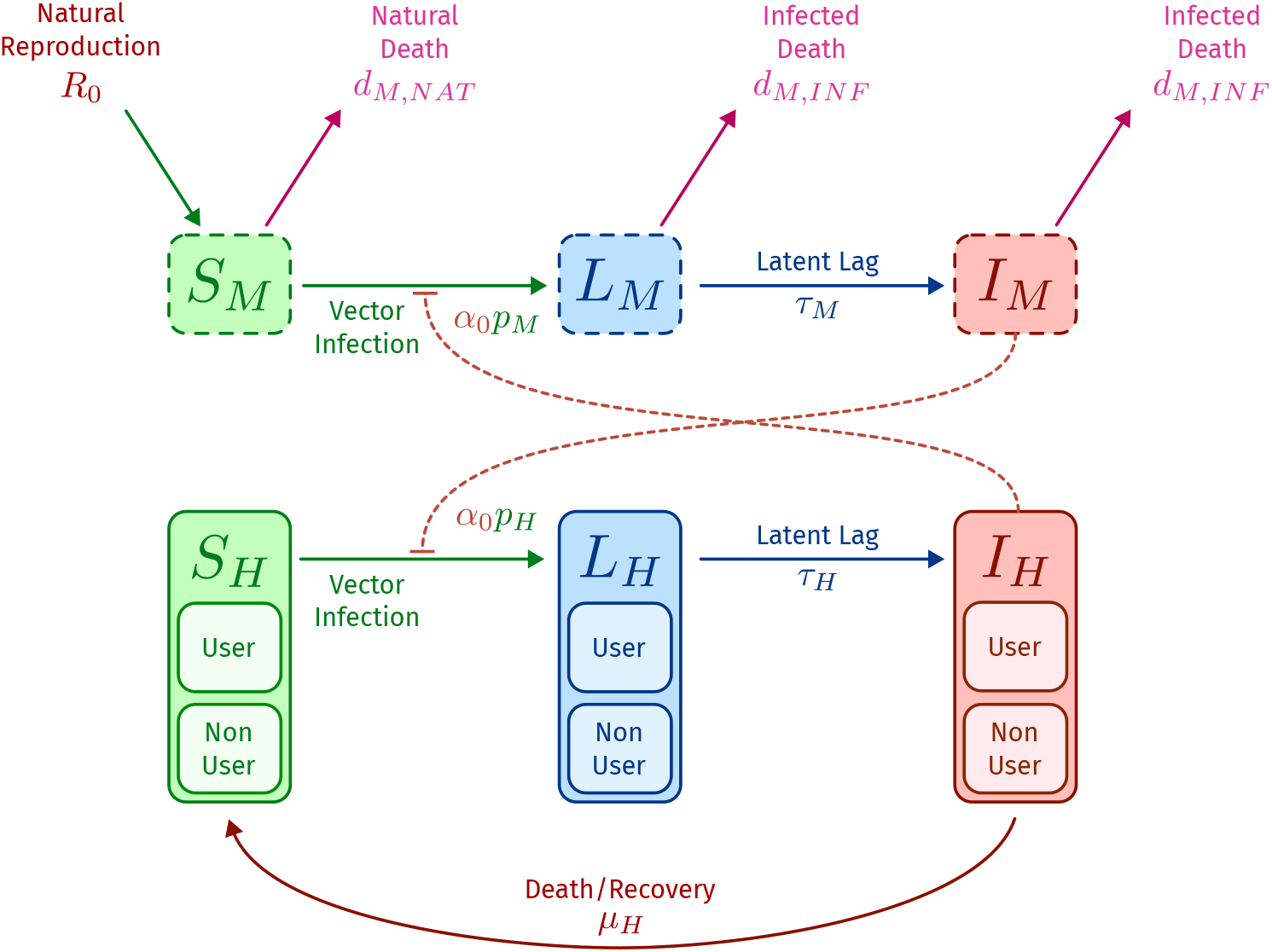
Schematic of the compartmental EIR model with delay including ITN intervention effects. Users and non-users are treated as separate components to describe different infection dynamics.

These are given by the following set of equations:

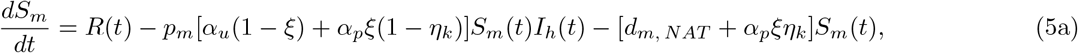

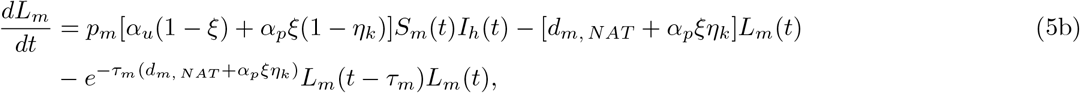

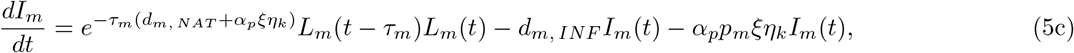

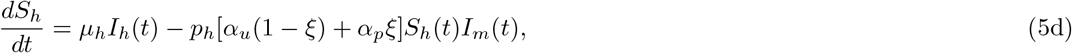

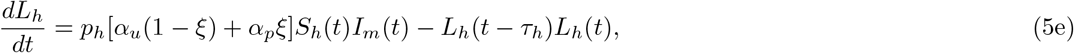

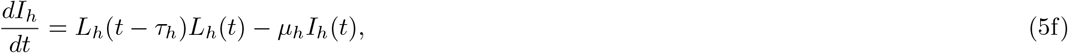

where human populations are separated into two cohorts – protected and unprotected – based on the proportion of ITN users given by *ξ*. These two cohorts differ in their experienced daily biting rate where

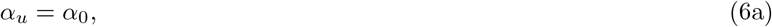

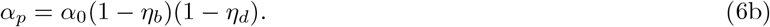

Like the non-intervention case, a state-state solution 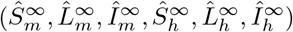 also exists for the case with ITN interventions. This can be numerically calculated as a function of model parameters Θ and intervention effects parameters *η*_*k*_, *η*_*d*_.*η*_*b*_, *ξ*.

### C. Efficacy-adjusted use

We define efficacy-adjusted use as the level of ITN use – consisting of fully efficacious, brand new nets – that would provide the same level of protection as that of the true scenario where ITN efficacy is compromised due to age and insecticide resistance. In this case, we take the term “protection” to be synonymous with the reduction in theoretical EIR calculated based on the above compartmental models conditioned on a set of chosen model parameters Θ. Therefore, the efficacy-adjusted use 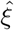 is calculated as a function of real use *ξ*, and ITN intervention effects *η*_*k*_, *η*_*d*_, *η*_*b*_ such that,

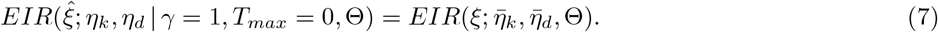

where 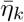, 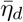 are weighted average of parameters based on the fraction of nets of type *i* and age *T* .

One important feature of this definition is the use of theoretical EIR as a normalising value to quantify the level of protection, rather than asserting them as equivalent to true EIR values which are often difficult to estimate and validate. By only allowing ITN related parameters (*ξ, η*_*k*_, *η*_*d*_, *η*_*b*_) to vary, this choice of normalisation affords robustness to the choice of biological model parameters Θ and isolates the effect of changing ITN use on protection.

## VI. MODEL EVALUATION AND VALIDATION

### A. Stability and robustness

Central to the our approach is a mechanistic model for mosquito-human transmission dynamics, which is subsequently used to calculate estimates of theoretical EIR. We emphasise that this estimate does not aim to represent the true absolute EIR, but instead captures the relative variations in EIR in the presence of interventions and environmental factors. In our analyses, we use biological model parameters to represent generic dynamical features such as transmission probabilities, latent infection times and death rates. Instead of re-calibrating model parameter values against experimental data, we adopt parameter values from results presented in existing data-driven and mathematical modelling studies. Since these parameter values are taken from studies that have been calibrated against observed datasets, their usage in the mechanistic model allows some degree of information from these datasets to be implicitly incorporated in our analyses. This approach achieves a balance between the data-requirements of the analysis, and mathematical flexibility of the model. Excluding latent period lengths *τ*_*m*_ and *τ*_*h*_ that vary in different environmental and disease settings, the base EIR model with no interventions utilises 6 parameters, which are listed alongside their value and source study in the Table V.

Because the efficacy of intervention methods are calculated based on a reduction in theoretical EIR, the effects of parameter misspecification for the purposes of calculated an efficacy-adjusted coverage is partially mitigated. However, this outcome is only applicable if the model estimates of theoretical EIR and trends in EIR reduction at different intervention levels are reasonably insensitive to variations in model parameter values. We perform three different tests to assess the robustness of the EIR model.

#### 1. Parameter sensitivity

To identify model parameters that pose the largest impact on EIR estimates, we first perform a simple sensitivity analysis for each parameter. To do this, each of the 6 model parameters are allowed to vary uniformly up to *±*50% of their original value and their relative impact on the steady state population proportions 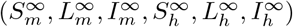 are observed. Variations are done for one model parameter at a time. For convenience, hat notation refers to quantities calculated for the case with perturbed parameter values. Therefore, relative impact on proportions calculated as 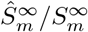 (similarly for other populations) represents the marginal effect of misspecification in a given parameter *θ* on the steady state solution. For each of the parameters tested, we run 1000 random perturbations from their base value and results are presented in Figure 4.

**FIG. 4.**
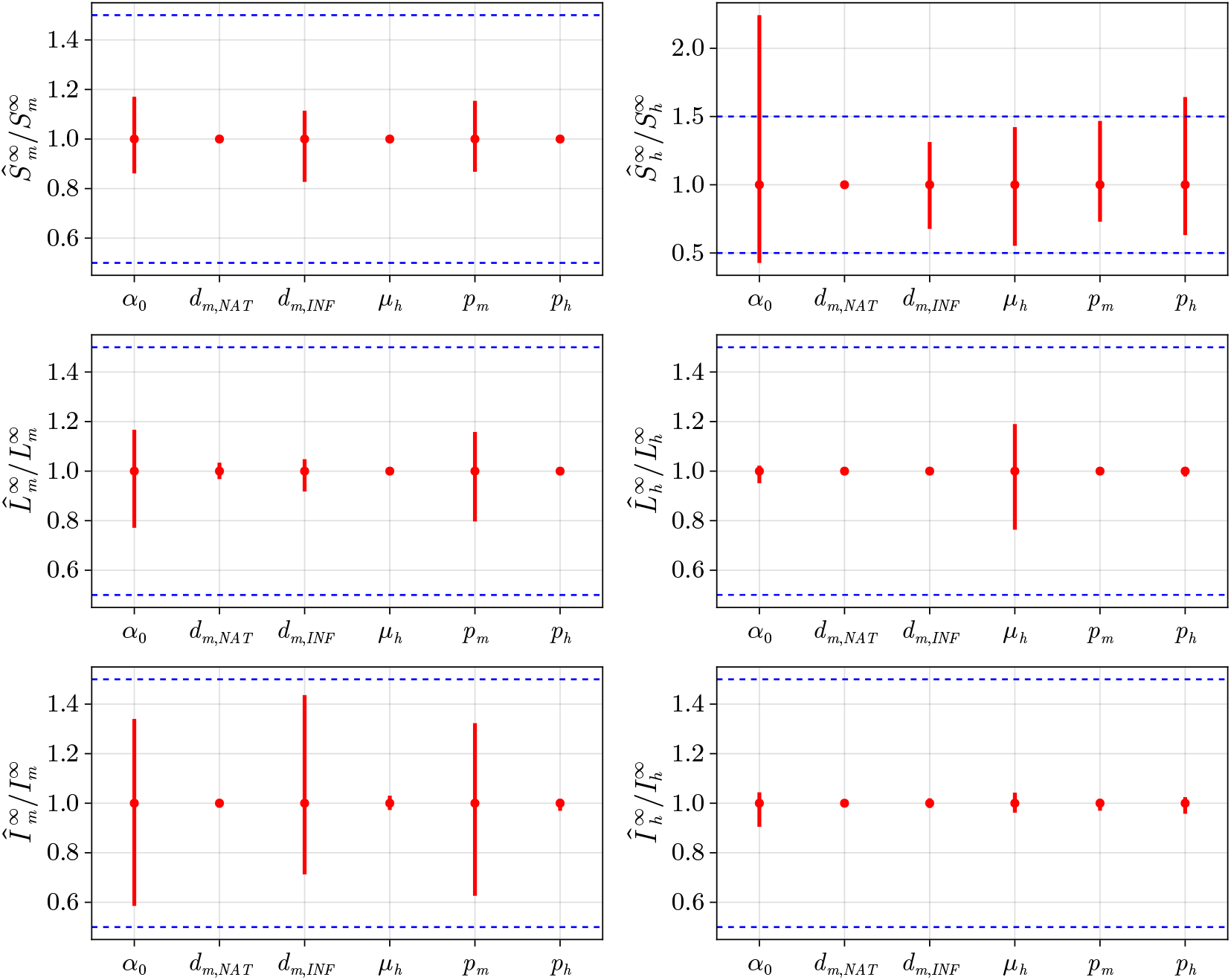
Sensitivity analysis of the base theoretical EIR model with no interventions. Each model parameter value is allowed to vary by *δ ∼*Uniform(*−*50%, 50%). Variation for each parameter is applied one at a time and other parameters are kept fixed. Dashed lines correspond to an equivalent *∼δ* Uniform(*−*50%, 50%) perturbation of steady state proportions using the baseline model parameter values. Columns that fall within dashed boundary lines indicate sub-linear sensitivity.

From the sensitivity analysis, we find that the marginal effects of parameter misspecification on final steady state proportions are sub-linear in all cases with the exception of the proportion of susceptible humans *S*_*h*_. An *m*% relative change in a given parameter value results in *< m*% change in steady state proportions. This suggests that mechanistic EIR model is generally robust to parameter misspecification. Of the parameters tested, the base human biting rate *α*_0_, infected mosquito death rate *d*_*m,INF*_ and probability of mosquito infection *p*_*m*_ have the largest impact on the steady state population of infected mosquitoes 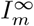, and thus theoretical EIR. This behaviour is unsurprising given that they are parameters the directly affect the frequency of infection opportunities for a given mosquito, and their rates of removal (i.e. due to death).

#### 2. Seasonal variation and human bite rates

Because the human bite rate *α*_0_ is one of the more influential parameters affecting theoretical EIR, it is important to test if these variations also impact EIR reduction ratios. That is, an idealised model of EIR should have reduction ratios that are stable across different EIR ranges. This behaviour should also be consistent across different levels of intervention coverage. The stability of reduction ratios is a favourable property as it allows for the direct comparison of scenarios across different epidemiological settings (e.g. seasonality, intervention composition).

To test the invariance in reduction ratios, we implicitly vary the base level of EIR (i.e. the non-intervention case) by applying a constant multiplier *c* on two different model parameters: the baseline mosquito density *N*_*m*,0_ and the human bite rate *α*_0_. The transmission reduction ratios are then calculated for varying levels of *c* and at different ITN uses with results given in Figure 5.

**FIG. 5.**
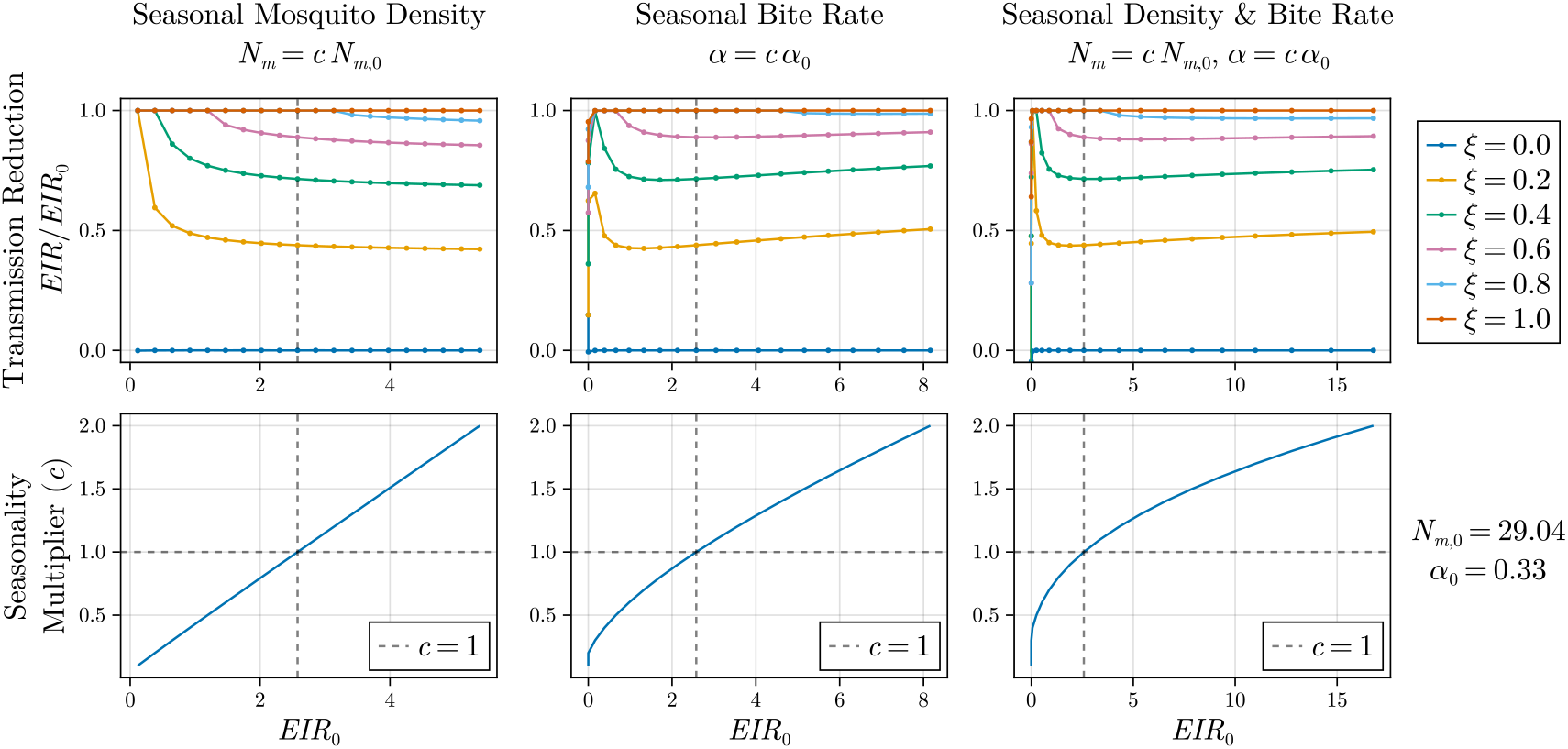
Plots of EIR reduction at various ITN use levels *ξ ∈ {*0, 0.2, 0.4, 0.6, 0.8, 1.0*}* and baseline *EIR*_0_ values. *EIR*_0_ values is mediated by applying a scaling factor *c ∈* [0, 2] to two model parameters: baseline mosquito density *N*_*m*,0_, and mosquito bite rate *α*_0_. Dotted lines correspond to the baseline reference of *c* = 1. Results show stabilty in EIR reduction rates across varying levels of *EIR*_0_.

In all cases with the exception of low EIR regimes, model estimates for transmission reduction is incredibly robust. These relationships are consistent across different levels of ITN coverage. This also suggests that theoretically derived EIR reduction ratios as a proxy for ITN efficacy is insensitive to parameter misspecification for the human bite rate *α*_0_.

#### 3. Misspecified death and infection rates

Two other parameters, the death rate of malaria infected mosquitoes *d*_*m,INF*_ and the probability of infection for Plasmodium susceptible mosquitoes per feeding attempt *p*_*m*_, are sources of significant sensitivity in steady state proportions and by extension, theoretical EIR estimates. We conduct similar analyses to that of Section VI A 1 but instead choose to vary *d*_*m,INF*_ and *p*_*m*_ and observe their effect of EIR reduction ratios at various ITN use levels, results shown in Figure 6. In all cases, whilst raw theoretical estimates of EIR varies with perturbations in the parameter, EIR reduction ratios remain relatively robust with the exception of very high infected mosquito death rates *d*_*m,INF*_ or low infection probability *p*_*m*,0_. In these extreme parameter regions, the scaling relationship between ITN use *ξ* and EIR reduction is most greatly affected for small values of *ξ*. This reaffirms the EIR model’s robustness to parameter misspecification when used to calculation reduction ratios.

**FIG. 6.**
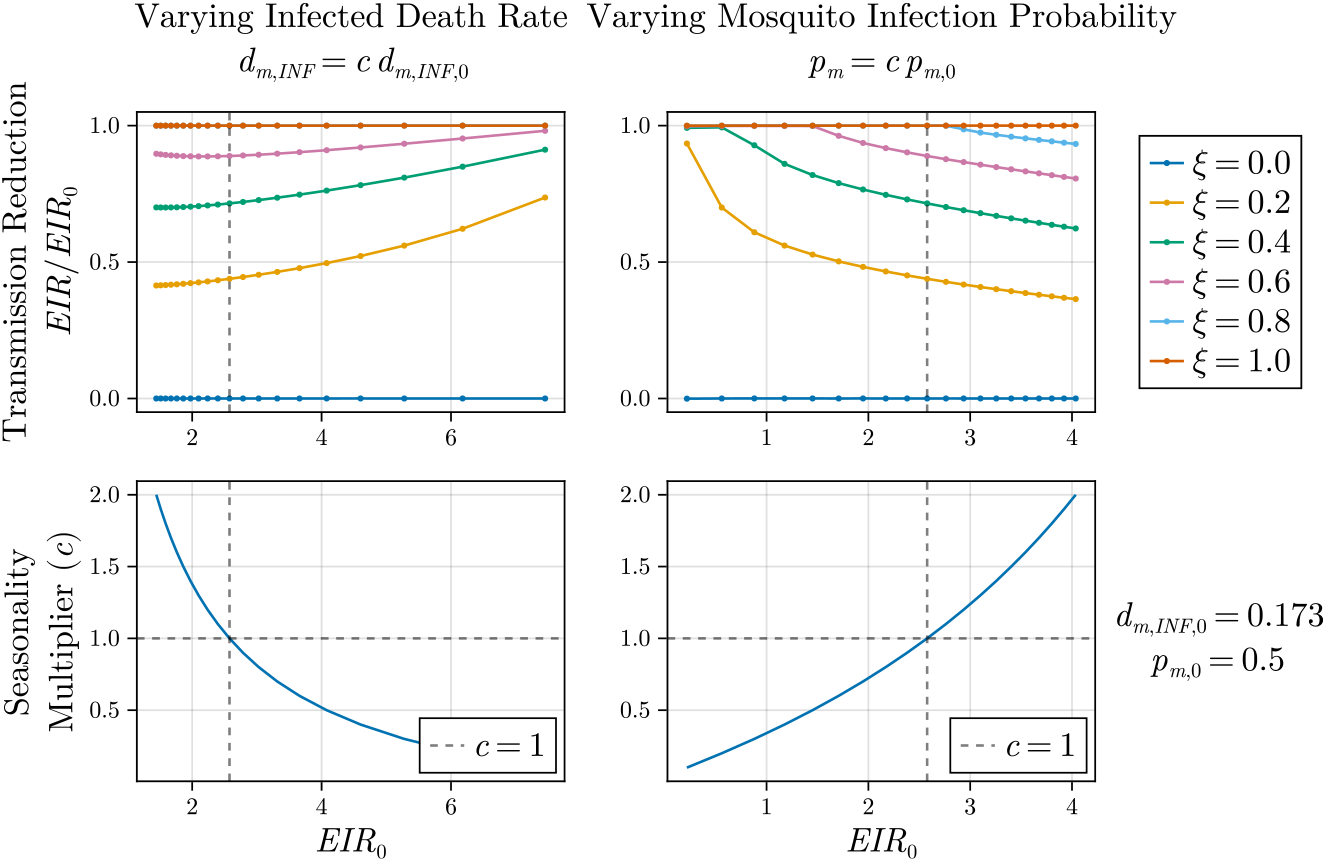
Plots of EIR reduction at various ITN use levels *ξ ∈ {*0, 0.2, 0.4, 0.6, 0.8, 1.0*}* and baseline *EIR*_0_ values. *EIR*_0_ values is mediated by applying a scaling factor *c ∈* [0, 2] to two model parameters: baseline infected mosquito death rate *d*_*m,INF*_ , and mosquito bite rate *p*_*m*_. Dotted lines correspond to the baseline reference of *c* = 1. Similar stability in the EIR reduction ratios are shown for moderate values of *c*. Trends become non-linear for either low rates of mosquito death *d*_*m,INF*_ or high rates of mosquito infection *p*_*m*,0_

### B. Comparison to reference model

The usage of bednets confers direct protection to users and indirect protection to non-users. Barrier effects confer an indirect protection by reducing the number infected humans, and thus the proportion of infectious mosquitoes. When insecticides are used (e.g. LLINs), an additional indirect benefit is attained due to the reduction in mosquito populations [20, 21].

The proposed EIR model with interventions does not include separated direct and indirect effects for ITNs when they are used. In Section IV C, the barrier effect parameter *η*_*b*_ is selected to best reproduce EIR reduction values for the scenario where only untreated nets are used to confer direct protection based on values provided by *Unwin et al*. [21]. To assess the consistency of the proposed EIR model against existing models, we test its ability reproduce the cumulative (i.e. both direct and indirect effects) EIR reduction ratios at different ITN use levels *ξ* presented by analysis by *Unwin et al*. To ensure a proper comparison, we assume that nets are held at a given use level *ξ* across a 3 year period and begin with initially full bioassay efficacy, and there are no insecticide resistance effects present. As nets age, we calculate the 3 year cumulative reduction in EIR and compare it against the case where no interventions are used. Comparisons of the EIR reduction ratios vs. ITN use *ξ* are shown in Figure 7.

**FIG. 7.**
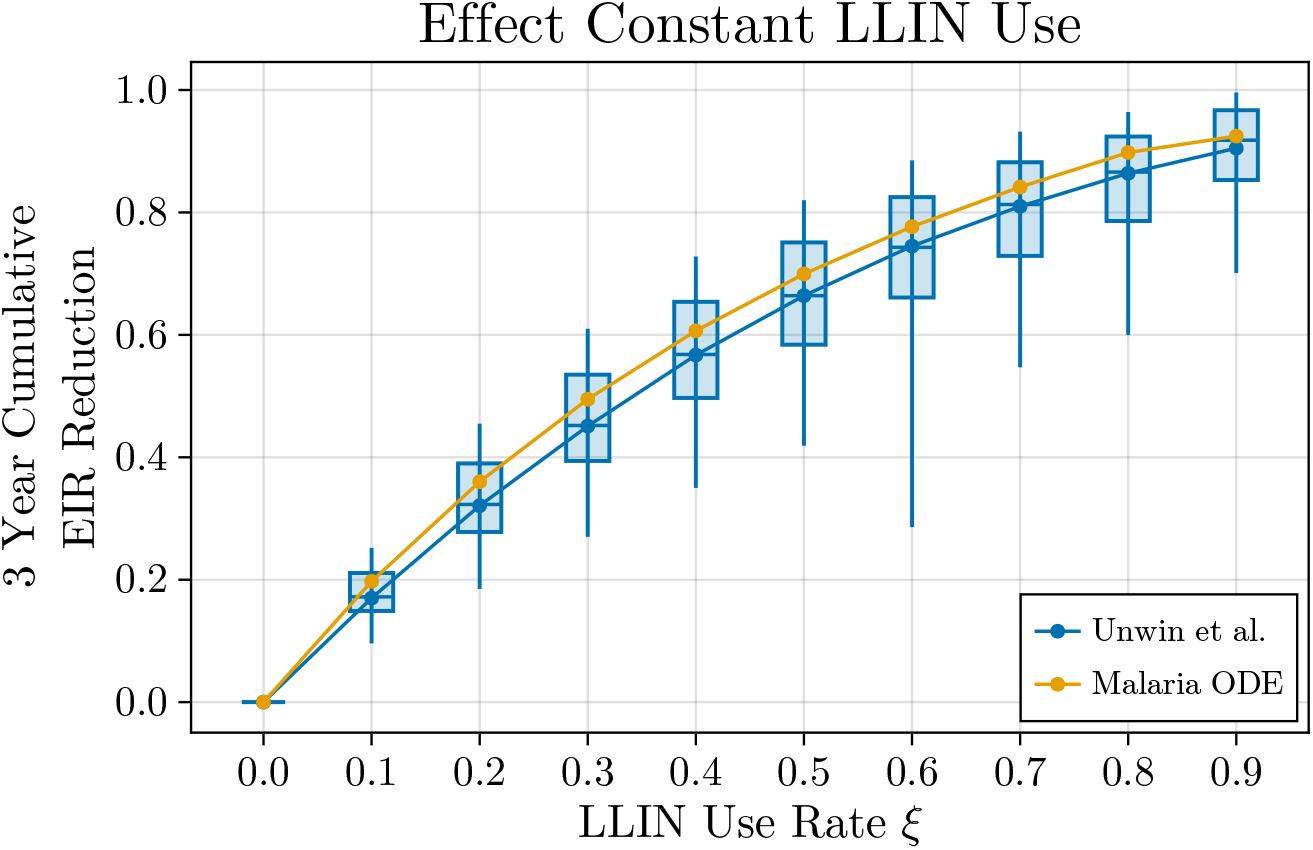
Total reduction in EIR including both direct user protection and indirect community protection effects as a function of LLIN use (*ξ*) for two different models: individual-based model by *Griffin et al*. and *Unwin et al*. [21] (blue), and proposed malaria EIR ODE model (yellow). Box and whiskers are taken directly from *Unwin et al*. where boxes correspond to the upper and lower quartiles, and whiskers are 1.5 IQR. The proposed simplified EIR model is able to closely reproduce the EIR Reduction-Use curves from more complex analyses.

Despite having different model formulations for EIR, we find that our proposed simplified EIR with added ITN effects is able to closely reproduce the nonlinear EIR reduction ratio curves reported by *Unwin et al*. Across the entire range of ITN use *ξ ∈* [0, 0.9], model estimates of EIR reduction values lie within the 1.5 IQR band estimates from model by *Unwin et al*. Furthermore, we find that the estimated value for barrier effects *η*_*b*_ = 0.323 is broadly in agreement with that of the data-calibrated estimates repellence probability for untreated nets (0.409) provided by *Unwin et al*. This close agreement not only affirms the findings from previous studies, but also assures that the design simplifications and components used in the EIR model do not detract from established findings of the existing models and literature.

## VII. RESULTS AND DISCUSSION

### A. Theoretical results

#### 1. Deterrence, killing and use relationships

To better understand the dynamics of the EIR model in response to killing, deterrence and use effects, we perform a full sweep of the parameter space and calculate the corresponding EIR reduction ratios. Results are shown in Figure 8. We find that the effect of deterrence *η*_*d*_ plays a large role in determining the scaling behaviour between ITN use *ξ* and conferred cumulative protection. When high levels of deterrence are present (i.e. *η*_*d*_ *→* 1), killing effects become redundant and protection scales linearly with ITN use *ξ*. This is expected as deterred mosquitoes never have the opportunity to make contact with ITN nets and thus do not get killed. This effect is equivalent to the case where bednets act as perfect barriers for protection. As the deterrence effect (*η*_*d*_) decreases, a nonlinear relationship emerges between the three parameters where both increasing values of killing *η*_*k*_ and use *ξ* contribute to the decrease in EIR. At extremely low ITN efficacy levels (i.e. *η*_*k*_ *→* 0, *η*_*d*_ *→* 0), the maximum attainable EIR reduction achieved is heavily impacted.

**FIG. 8.**
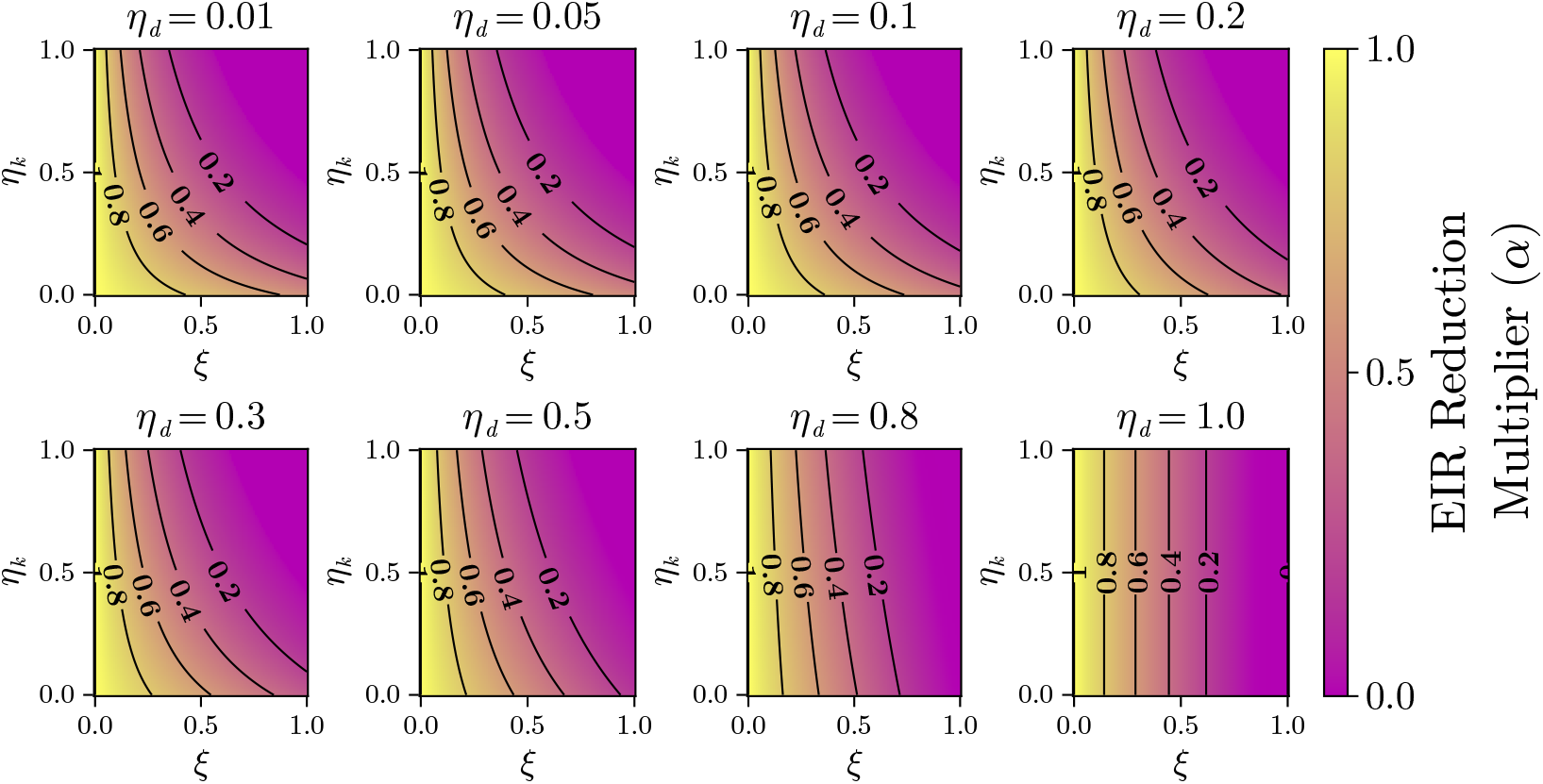
Phase plot of the predicted reduction multiplier in theoretical EIR due to ITN interventions at at various levels of killing (*η*_*k*_), deterrence (*η*_*d*_) and use (*ξ*). A barrier effect of *η*_*b*_ = 0.372 is applied in all cases. A multiplier value of 0 indicates a 100% reduction in EIR (i.e. perfect protection) and higher values indicate less protection. Nonlinear behaviours are observed in the low deterrence regime where both killing effects (*η*_*k*_) and ITN use (*ξ*) both contribute to EIR reduction. For high deterrence *η*_*d*_ *>* 0.8, the importance of killing effects is diminished and protection is proportional to ITN use.

#### 2. Age effects

In practice, the effect of killing *η*_*k*_ and deterrence *η*_*d*_ are highly correlated with each other. This is further complicated by the fact that not all users possess nets of equal age and thus effectiveness at any given time. This complex relationship can be described by combining a model of waning bioassay efficacy, statistical models for converting bioassay efficacy to in-field deterrence and killing effects [31] (see Section IV B and Supplementary Information) and measures of net age. This can be used together with Equation 7 to calculate a value for efficacy-adjusted net use *ξ*^*\**^ defined as the use level of nets that are undegraded and fully efficacious (bioassay mortality) that would provide an equivalent level of EIR reduction. The relationship between real use *ξ*, net age *T*_*age*_ and efficacy-adjusted use *ξ*^*\**^ is shown in Figure 9.

**FIG. 9.**
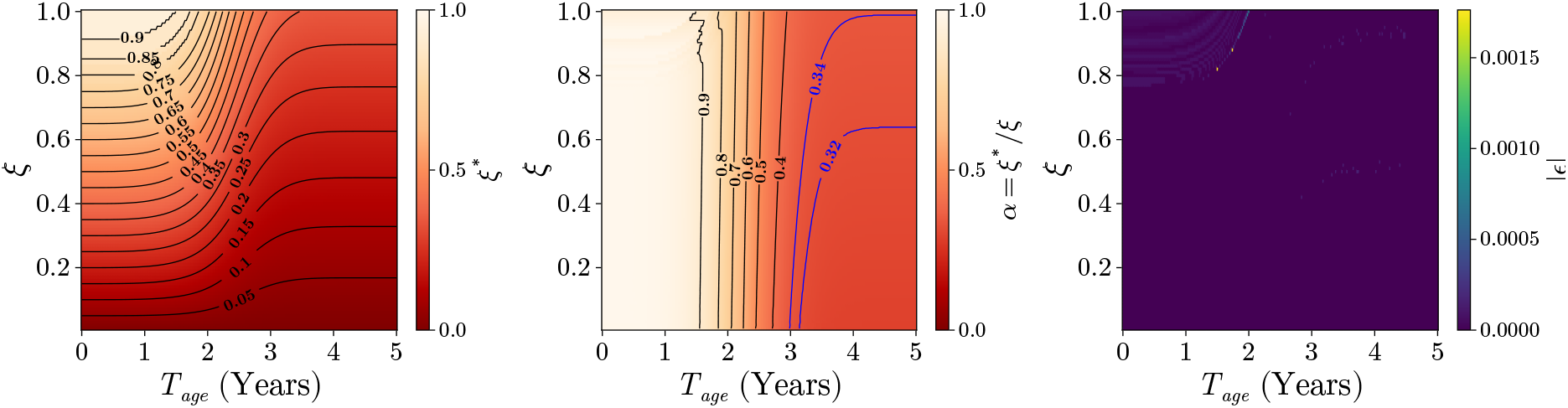
Plots of efficacy-adjusted use as a function of raw ITN use and average net age. Calculation of theoretical EIR fully incorporates the bioassay mortality to in-field deterrent and killing relationship proposed by *Nash et al*. [31], and the fitted waning bioefficacy Weibull model for age effects. From left to right: (1) isoclines of efficacy-adjusted use (*ξ*^*\**^) as a function of raw ITN use (*ξ*) and average net age (*T*_*age*_) with a critical transition at *T*_*age*_ *≈*2.5 years. (2) Proportional reduction in protection (*α* = *ξ*^*\**^*/ξ*) as a function of raw ITN use (*ξ*) and average net age (*T*_*age*_). (3) Errors from solving Equation 7 showing excellent fits across the whole domain. generally retain their full level of bioefficacy and thus variations in use *ξ* closely align with equal variations in efficacy-adjusted use *ξ*^*\**^. A significant nonlinearity occurs for intermediate mean net age (1-3 years) with a critical transition occurring around mean net age of 2.5 years where the gap between raw use and efficacy-adjusted use (*ξ − ξ*^*\**^) rapidly widens for every small increase in mean net age and the maximum attainable EIR reduction starts to decrease. Beyond a mean net age of 3 years, a perfect level of real ITN use *ξ* = 1 will only yield equivalent protection to that of use level *ξ*^*\**^ = 0.43.

Steady-state solutions for Equation 5 are found numerically using a bisection algorithm with an early break condition when errors between EIR values of the true and ideal case are less than 10^*−*4^. Throughout the entire domain of values, an excellent level of fit is attained as shown in the error plot of Figure 9. For lower mean net ages (*<* 1 year), ITNs

### B. Continental trends and counterfactual analysis

The proposed EIR reduction ratios can be combined with raw ITN use outputs from the MITN model to calculate spatial maps [4] of efficacy-adjusted use. These high-resolution spatial estimates extend previous measures of ITN use to account for efficacy penalty effects such as from insecticide resistance or ageing net crop. Similar to the analyses by *Tan et al*., spatiotemporal rasters for efficacy-adjusted use can be subsequently used to calculate aggregate statistics of key ITN coverage measures at multiple spatial scales (e.g. national, subnational, district).

We follow the methodology and data sources use in *Tan et al*. and calibrate an MITN model with surveys across 44 countries during the period of 2006-2024. Pixel level use is combined with subnational (Admin 1) level estimates of net age composition and type to calculate pixel level estimates of efficacy-adjusted use. Aggregate measures of efficacy-adjusted use are then calculated as a population weighted average using pixel level population estimates taken from IHME datasets. Comparisons of raw and efficacy-adjusted use across 2006-2024 are shown alongside continent level aggregates with each counterfactual in Figure 11. Country level adjusted use is given in the Supplementary Information.

**FIG. 11.**
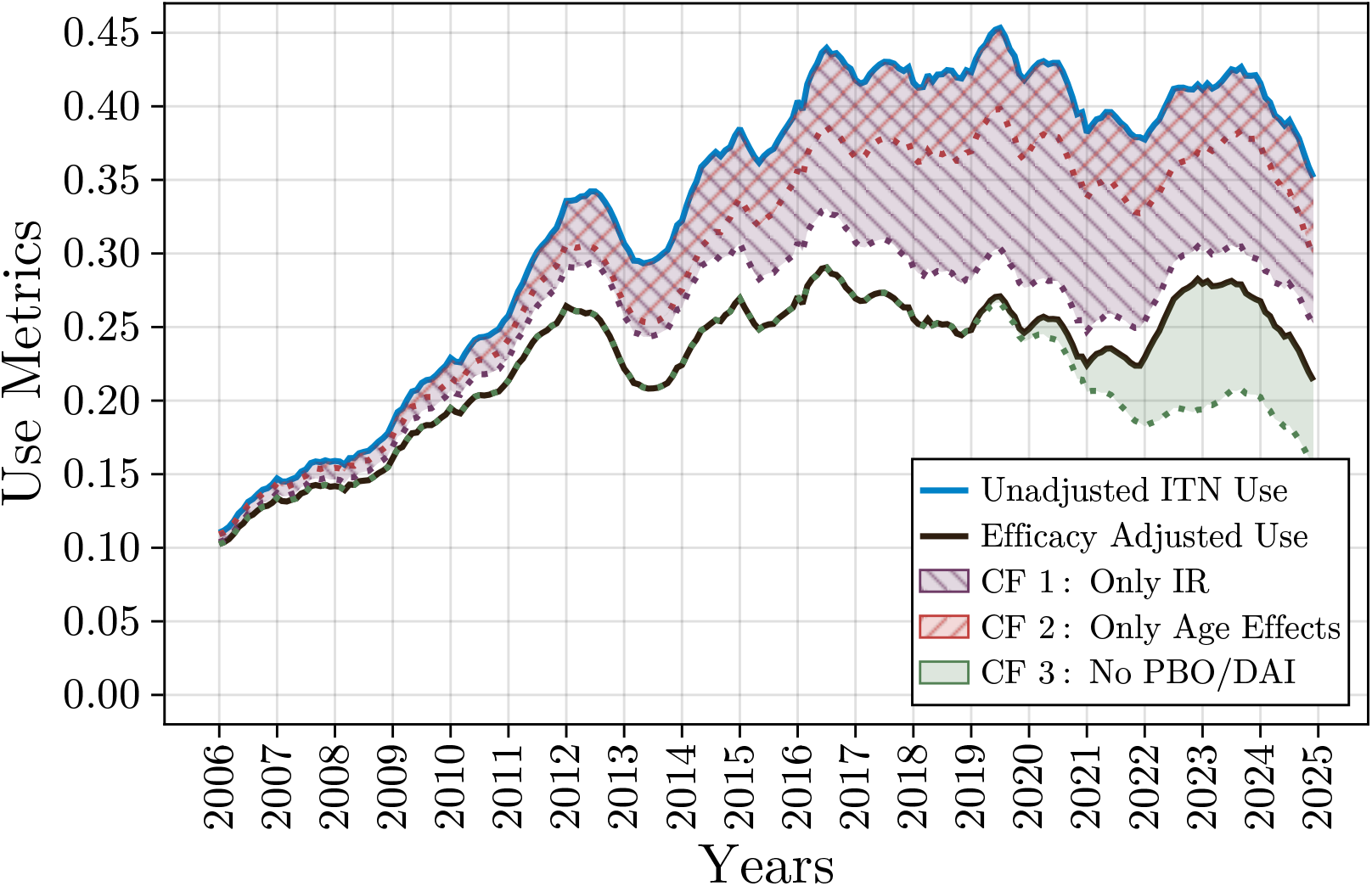
Raw ITN use, and efficacy-adjusted use for three different counterfactuals: (CF1) No age related penalties, (CF2) No insecticide resistance, and (CF3) no deployment of next-generation LLINs. Shaded regions indicate the level of penalty relative to the raw ITN use for each counterfactual. Shaded green region describes the amount of ITN protection loss that is mitigated due to the adoption of more efficacious next-generation LLINs (e.g. PBO and DAIs). Hashed regions indicate the marginal effects of two different phenomenon (age and IR). Large growth in disparity from 2010-2017 is present. Disparity stabilises from 2017 accompanied for a stagnation in ITN use.

In addition to estimates of current efficacy-adjusted use, we also analyse three different counterfactual scenarios: (1) Only insecticide resistance effects present (purple hashed), (2) only waning bioefficacy due to age effects present (red hashed), and (3) no distribution of next generation LLINs (i.e. PBO and DAI) (green shaded). To simulate Scenario 1 where we examine the impact of having no age related degradation, an upper limit on the allowable net age *T*_*max*_ is imposed on the net crop and thus guaranteeing that the mean net age is always young. The remaining fraction of nets that exceed this age based on the net age composition output by the MITN are redistributed into the age bins below the *T*_*max*_ according to the pre-existing proportions. We consider various maximum age values of *T*_*max*_ = *{*6, 12, 24, 36*}* months (see Figure 10). Scenario 2 is applied by setting the baseline vector susceptibility to insecticide as *γ* = 1 regardless of net type. Finally, Scenario 3 is tested by calculating *v*(*t*) with the assumption that the entire net crop at any given time is composed of only cITNs and LLINs, and any distributed volumes of PBO and DAI being reallocated to LLINs.

**FIG. 10.**
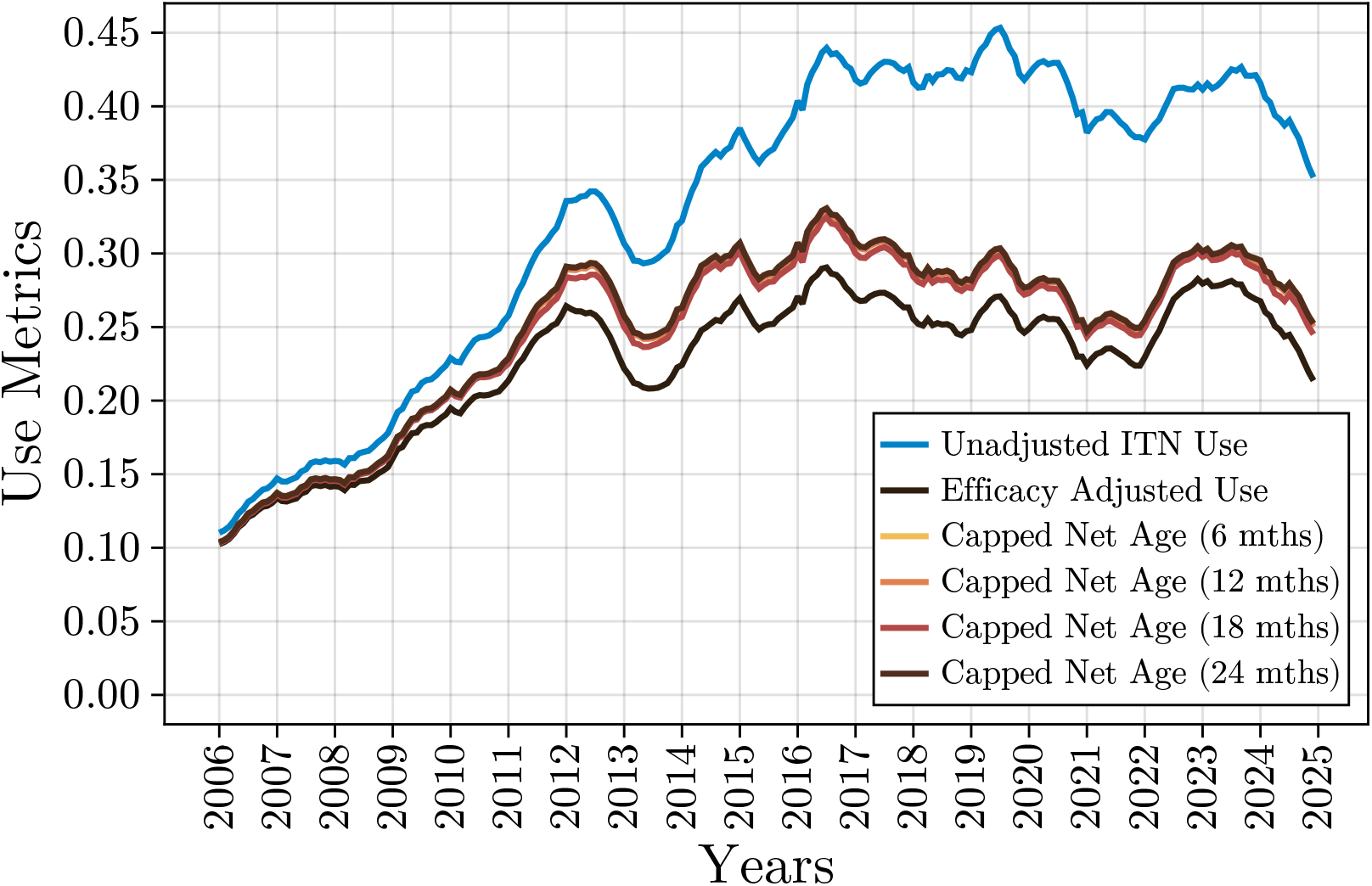
Population weighted average raw ITN use, and efficacy-adjusted use with average net age (*T*_*age*_) capped at various thresholds for the African continent.

Broadly, Scenario 1 and 2 represent counterfactual cases where either age effects or insecticide resistance (IR) are absent respectively. They can be used in combination to test the independence between IR and age related penalties with respect to the proposed mode EIR. If these effects are independent, they allow the proportional attribution of estimated penalties in ITN use Δ_*ξ*_ = *ξ − ξ*^*\**^ to IR and age, and thus identify the driving factors of reduced intervention efficacy in different regions. Finally, the counterfactual Scenario 3 represents the case where no next generation LLINs were adopted to combat IR effects can be compared against the actual efficacy-adjusted use to quantify the estimated Unadjusted ITN Use Efficacy Adjusted Use Capped Net Age (6 mths) Capped Net Age (12 mths) Capped Net Age (18 mths) Capped Net Age (24 mths) impact of the adoption of next generation LLINs. Plots of the true and counterfactual aggregate estimates of efficacy-adjusted use for the continental level are given in Figure 11, and at country level (see Supplementary Information).

On a continental level, we find that there is a significant growing disparity between raw estimates of ITN use and those after accounting for deleterious effects such as insecticide resistance and age-related degradation. In 2010, this disparity was approximately *ξ − ξ*^*\**^ = 0.03 amounting to a penalty rate of 15% from the raw ITN use estimate of 0.23, which is equivalent to an efficacy of 85% (i.e. the equivalent level of ITN use accounting for efficacy is 85% of the raw ITN use). A decade later in 2020, we estimate a large growth in this disparity to *ξ − ξ*^*\**^ = 0.15 equivalent to a penalty rate of 39% from the raw ITN estimate. Despite falling ITN use from 2020 to 2024, this trend in the disparity between raw and adjusted use has stabilised from 2020. However, disparities remain significant with the most current estimates at the end of 2024 show an efficacy-adjusted use of 0.21, lower than the the naive raw use estimate of 0.35. To decompose this observed disparity, we first test the independence of insecticide resistance and age effects. To do this, we compare the true efficacy-adjusted use estimates against the product of the marginal efficacy-adjusted use containing either insecticide resistance or age effects in isolation (i.e. Scenario 1 and 2). From Figure (see Figure 12), we see a strong agreement in both values. This indicates that conditioned on the proposed EIR model, insecticide resistance and age effects can be treated as independent. The proportion of penalty that can be attributed to each effect (insecticide resistance or age) can thus be calculated as follows,

**FIG. 12.**
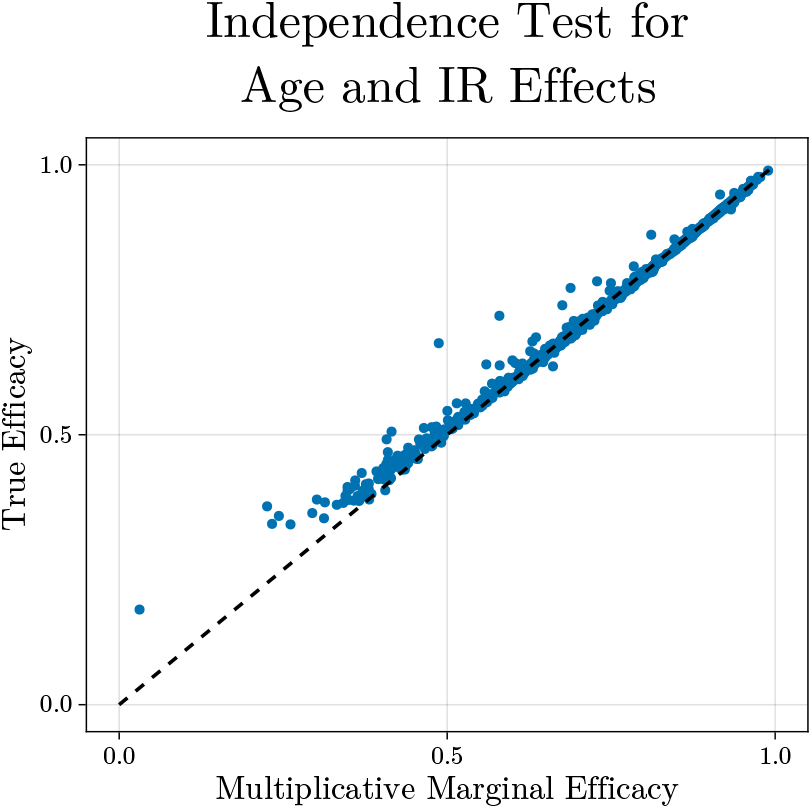
Plots of the true efficacy (*ξ*^*\**^*/ξ*) and the multiplicative marginal efficacy taken as the product of 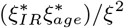. Closeness to the main diagonal indicates strong independence between both age and insecticide resistance effects in the model.

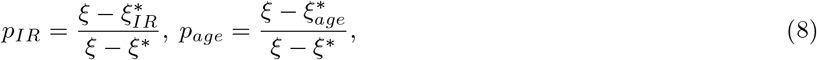

where 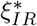 and 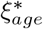 are the efficacy-adjusted use assuming only insecticide resistance (Scenario 2) or age effects (Scenario 1) are present respectively. On a continental level, we find the proportion of the disparity between raw and adjusted use that is attributable to insecticide resistance effects has increased substantially from 2010 with fastest period of growth occurring between 2010-2016. This is in agreement with analyses by [7] that identified increasing trends in insecticide resistance in the African region throughout 2005-2017. In earlier years prior to 2012, both insecticide resistance and age effects played comparable but minor roles in reducing the efficacy adjusted use. This is likely driven by the lower prevalence of insecticide resistance and ramping up of ITN distribution campaigns during this period, corresponding to younger net crop. As of 2024, we estimate that 72% of the reduction in actual protection is attributed to insecticide resistance.

The impact of insecticide resistance is already well recognised and has driven the recent increased adoption of next generation LLINs. In the last 5 years, these adoption trends have been led by countries such as Liberia, Zambia and

Mozambique (see Figure 13). It is estimated that 34% of total nets currently in use in Africa are next generation LLINs (i.e. PBO or DAI) as of 2024 [4]. By comparing current efficacy-adjusted use estimates against the counterfactual where no next generation LLINs are distributed (Scenario 3), one can calculate the amount of protection penalty that has been mitigated due to next generation LLIN adoption (see Figure 11). We estimate that the use of next generation LLINs has averted a potential further reduction of 16% in the efficacy-adjusted use, where the counterfactual scenario with no next-generation LLINs provide and equivalent protection to a ITN use of 0.16. At a country level, countries with the highest levels of insecticide resistance were the greatest benefactors from next generation LLIN adoption. Unsurprisingly, countries that have adopted next-generation LLINs (PBOs & DAIs) in their distribution programs have generally been successful at maintaining the efficacy of their ITN interventions. (see Figure 13). However, the story is more complex when viewed across the most recent six years of history (2018-2024). During this period, improved ITN efficacy is not necessarily accompanied by improved use.

**FIG. 13.**
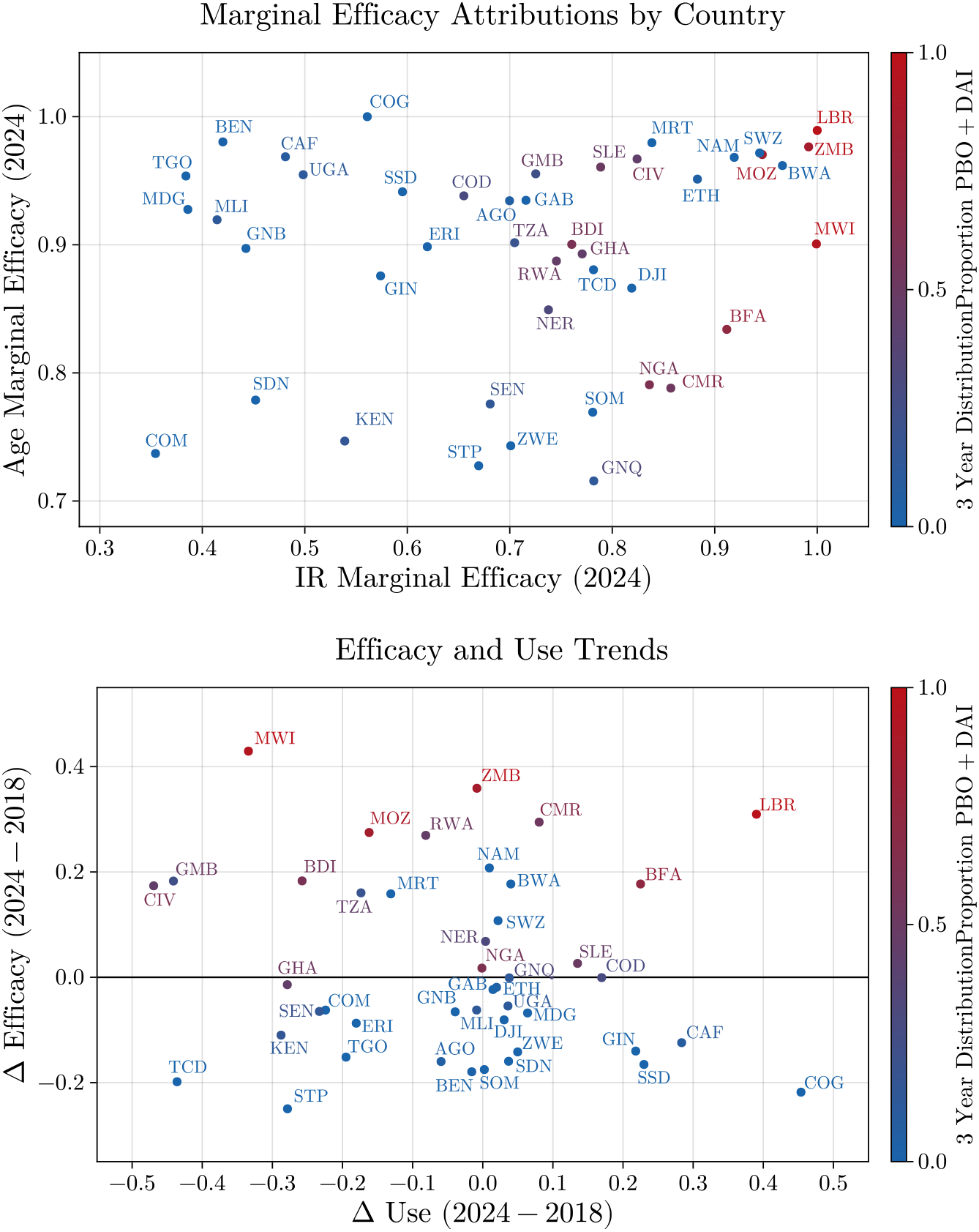
Breakdown of efficacy and use effects for 44 modelled countries based on proportion of next-generation LLINs distributed in the most recent 3 years (2022-2004). Top: Breakdown of efficacy penalties by age and insecticide resistance (IR) effects. The magnitude of IR related penalties generally larger than age related penalties. Negative values indicate a decrease in the efficacy and use values and thus compromised protection. Bottom: Comparison of change in use and efficacy between 2018 and 2024. Countries with a higher proportion of next-generation LLINs preserve a higher level of efficacy. However, this does not necessarily equate to improved overall protection improved efficacy is not correlated with improved rates of ITN use.

A spectrum of behaviour is present, where some countries have improved their use but suffer a penalty of decreased efficacy due to increased insecticide resistance and age effects (e.g. Congo, Guinea). Conversely, there are also countries with high ITN net crop diversity with high efficacy but have experienced declining levels of ITN use (e.g. Mozambique, Tanzania) thus resulting in overall lower real protection. This highlights the heterogeneous impact of ITN efficacy penalties such as insecticide resistance and age effects on countries, which is further confounded with varying levels of ITN use.

## VIII. CONCLUSIONS

In this paper, we have presented a novel method of calculating an alternative metric of ITN coverage, efficacy-adjusted use, to quantify the effective level of malaria protection afforded by a given ITN use. This metric combines information on insecticide resistance, net age and type composition with a proposed surrogate model of EIR to produce a more nuanced estimate of protection that accounts for factors that reduce ITN efficacy in the field. We emphasise that our analyses is only limited to the study of insecticide resistance (IR) and age effects on protection, and does not account for other extraneous factors.

The proposed EIR model is inspired from various compartmental modelling approaches by *Griffin et al*. [38, 39] and is used to calculate EIR reduction ratios in the presence of an intervention, which is subsequently used as a proxy for coverage. Notably, we innovate on previous well established models by prioritising parsimony in the model design.

The proposed model is greatly simplified and utilises a small number of 6 parameters with all but 3 requiring direct calibration against data. This is in comparison to the far more descriptive and complex state-of-the-art models that utilise upwards of 30 parameters, Despite this large simplification, we demonstrate that this model can be readily solved for steady states, and find that it is sufficiently complex to reproduce baseline EIR reduction curves in response to general LLIN use that were previously predicted by *Nash et al*. [31] (see Figure 7). Importantly, the proposed EIR model separately describes critical features such as age related degradation in bioefficacy and insecticide resistance.

This enables the EIR reduction – and by proxy the level of protection – to be estimated for any ITN use level, and net crop of any age and type composition.

We demonstrate the utility of the proposed approach by applying it to spatiotemporal estimates of ITN use, which is provided by the MITN model proposed by *Tan et al*. [4, 13] for modelling ITN coverage. Previous studies have noted that ITN coverage has lagged since its rapid growth in 2015, a trend that is consistent across Africa with the latest continent levels of ITN use being 0.36. Accounting for net age and insecticide resistance related factors, we find that the effective level of protection may be much lower and is approximately equivalent to an ITN use of 0.25 based on the reference case where all ITNs are maximally efficacious. This notion of equivalent level of ITN use, which we term efficacy-adjusted use, has a disparity from estimated raw ITN use that has steadily increased since 2010 and stabilised from 2020.

Two contributing factors for this penalty in protection are studied: insecticide resistance effects and age degradation effects. We find that age related penalties become more significant when net ages exceed 2 years (see Figure 8) and aligns with existing findings where the functional life time of nets is substantially less than the WHO recommended 3 years [47]. However, age related contributions are dwarfed by the effect of growing insecticide resistance and accounts for up to 65% of the estimated decrease in actual protection. On a country level, this disparity is largest in countries with large levels of insecticide resistance such as Madagascar, Benin and Comoros (see Figure 13).

A counterfactual scenario where no next generation LLINs were distributed to assess the efficacy of recent PBO and DAI distribution campaigns are also explored. Assuming that these new nets are fully bioefficacious, we unsurprisingly find that effective protection levels would be lower if new net types were not adopted. However, the magnitude of this decrease is highly significant where the adoption of next generation nets has potentially mitigated a further 16% reduction in protection. This amounts to an efficacy-adjusted use of 0.21, in comparison to the counterfactual value of 0.16. These results provides strong broad level support for the adoption of next generation LLINs to tackle the growing threat of insecticide resistance in the short-to-medium term.

The findings on efficacy-adjusted use also highlights key points of consideration from a modelling perspective. Net use, nets per capita (NPC) and net access are critical measures that act as baseline descriptors of ITN coverage. These measures, and in particular net use, are frequent inputs in many downstream modelling and statistical analyses to capture the effects of ITN intervention on epidemiological outcomes such as parasite rates, disease prevalence and incidence. However, in its raw form ITN use does not capture the in-field impact of nets on critical processes like rates of infection, and also do not account for heterogeneities in ITN performance due to availability of different levels of age related degradation and net types. The large disparity between an estimated in-field level of protection compared to raw values of ITN use suggest that there is a potential consistent overestimate in the level of effective protection afforded by ITNs. Furthermore, we find that the distribution of next-generation LLINs to maintain ITN efficacy has not always been accompanied with a sustained level of ITN use. This may result in effective levels of protection that are not necessarily improved despite the adoption of newer, more bioefficacious ITNs.

In summary, our analyses offer a mechanistically-informed insight on the potential mismatch between unadjusted ITN use and those accounting for deleterious effects on ITN effectiveness. We attempt to consolidate modelling results across various aspects of ITN interventions (coverage, bioefficacy, durability, insecticide resistance) to provide an informed initial estimate of the degree of mismatch. This highlights the importance of considering a notion of efficacy-adjusted use to better represent the level of protection afforded by ITNs and its associated spatiotemporal heterogeneities. We note that our analyses inherit similar limitations to its component models, and provide an avenue for further research and refinement. These include the inclusion of the effect of private sector in household ITN acquisition to provide more accurate estimates of net age and type, further refinements and validation on the estimates of epidemiological and biological parameters of the EIR model, and exploring the relationship between net age and net retention in households. Recently consolidated databases of field and lab studies such as those by CASTAnet may also provide further refinements of models of longitudinal ITN bioefficacy and durability.

## Data Availability

All data produced in the present work are available upon reasonable request to the authors. Code and implementations are accessible at the following repositories:
ODE Model:
https://github.com/eugenetkj98/MalariaODE
MITN Model:
https://github.com/eugenetkj98/MITN-Public-v2-DEMO

https://github.com/eugenetkj98/MITN-Public-v2-DEMO

https://dhsprogram.com/

https://mics.unicef.org/

https://www.malariasurveys.org

https://allianceformalariaprevention.com/working-groups/net-mapping/

## ACKNOWLEDGMENTS

This work was supported, in whole or in part, by the Gates Foundation [INV-055192]. The conclusions and opinions expressed in this work are those of the author(s) alone and shall not be attributed to the Foundation. Under the grant conditions of the Foundation, a Creative Commons Attribution 4.0 License has already been assigned to the Author Accepted Manuscript version that might arise from this submission. This work also includes funding support from the Australian Government, National Health and Medical Research Council (Award No: GNT2025280). N.G. is supported by the Stan Perron Charitable Foundation and an NHMRC Investigator Grant (2041810).

## References

[1] W. H. Organization, World malaria report 2024 (World Health Organization, 2022).

[2] D. J. Weiss, P. A. Dzianach, A. Saddler, J. Lubinda, A. Browne, M. McPhail, S. F. Rumisha, F. Sanna, Y. Gelaw, J. B. Kiss, et al., Mapping the global prevalence, incidence, and mortality of Plasmodium falciparum and Plasmodium vivax malaria, 2000–22: a spatial and temporal modelling study, The Lancet 405, 979 (2025).

[3] S. Bhatt, D. J. Weiss, E. Cameron, D. Bisanzio, B. Mappin, U. Dalrymple, K. Battle, C. L. Moyes, A. Henry, P. A. Eckhoff, et al., The effect of malaria control on plasmodium falciparum in Africa between 2000 and 2015, Nature 526, 207 (2015).

[4] E. Tan, M. van den Berg, A. Saddler, C. Vargas, D. J. Weiss, A. Bertozzi-Villa, S. Bhatt, T. L. Symons, and P. W. Gething, Insecticide-treated bednet coverage in Africa 2006-2024: a spatiotemporal analysis of net ownership, use, age and type, medRxiv , 2025 (2025).

[5] H. Koenker, J. Yukich, M. Erskine, R. Opoku, E. Sternberg, and A. Kilian, How many mosquito nets are needed to maintain universal coverage: an update, Malaria Journal 22, 200 (2023).

[6] W. H. Organization, Global technical strategy for malaria 2016-2030 (World Health Organization, 2015).

[7] P. A. Hancock, C. J. Hendriks, J.-A. Tangena, H. Gibson, J. Hemingway, M. Coleman, P. W. Gething, E. Cameron,S. Bhatt, and C. L. Moyes, Mapping trends in insecticide resistance phenotypes in african malaria vectors, PLoS Biology 18, e3000633 (2020).

[8] S. W. Lindsay, M. B. Thomas, and I. Kleinschmidt, Threats to the effectiveness of insecticide-treated bednets for malaria control: thinking beyond insecticide resistance, The Lancet Global Health 9, e1325 (2021).

[9] N. Golding, Technical summary: Insecticide resistance space-time cube, Unpublished (2022).

[10] N. Golding, The rise of insecticide resistance in malaria vectors across Africa 2000-2025, Personal communications (2025).

[11] A. Bertozzi-Villa, C. A. Bever, H. Koenker, D. J. Weiss, C. Vargas-Ruiz, A. K. Nandi, H. S. Gibson, J. Harris, K. E. Battle, S. F. Rumisha, et al., Maps and metrics of insecticide-treated net access, use, and nets-per-capita in Africa from 2000-2020, Nature Communications 12, 3589 (2021).

[12] S. Bhatt, D. J. Weiss, B. Mappin, U. Dalrymple, E. Cameron, D. Bisanzio, D. L. Smith, C. L. Moyes, A. J. Tatem,M. Lynch, et al., Coverage and system efficiencies of insecticide-treated nets in africa from 2000 to 2017, Elife 4, e09672 (2015).

[13] E. Tan, M. van den Berg, C. Vargas, P. W. Gething, and T. L. Symons, Dynamical and time series approach to understanding compartmental stock and flow models: a case study in malaria intervention models, Journal of the Royal Society Interface (2026).

[14] O. O. Diallo, A. Diallo, K. B. Toh, N. Diakité, M. Dioubaté, M. Runge, T. Symons, E. M. Diallo, J. Gerardin, B. Galatas, et al., Subnational tailoring of malaria interventions to prioritize the malaria response in Guinea, Malaria Journal 24, 62 (2025).

[15] W. H. Organization, Guidelines for Malaria (World Health Organization, 2025).

[16] T. L. Symons, J. Lubinda, M. McPhail, A. Saddler, M. van den Berg, H. Baggen, Y. Berman, S. Hafsia, R. Jayaseelen, P. Amratia, et al., Estimating the potential malaria morbidity and mortality avertable by the US President’s Malaria Initiative in 2025: a geospatial modelling analysis, The Lancet 405, 2231 (2025).

[17] T. L. Symons, A. Moran, A. Balzarolo, C. Vargas, M. Robertson, J. Lubinda, A. Saddler, M. McPhail, J. Harris, J. Rozier,et al., Projected impacts of climate change on malaria in Africa, Nature , 1 (2026).

[18] G. F. Killeen, T. A. Smith, H. M. Ferguson, H. Mshinda, S. Abdulla, C. Lengeler, and S. P. Kachur, Preventing childhood malaria in africa by protecting adults from mosquitoes with insecticide-treated nets, PLoS Medicine 4, e229 (2007).

[19] S. S. Lim, N. Fullman, A. Stokes, N. Ravishankar, F. Masiye, C. J. Murray, and E. Gakidou, Net benefits: a multicountry analysis of observational data examining associations between insecticide-treated mosquito nets and health outcomes, PLoS Medicine 8, e1001091 (2011).

[20] G. F. Killeen, Control of malaria vectors and management of insecticide resistance through universal coverage with next-generation insecticide-treated nets, The Lancet 395, 1394 (2020).

[21] H. J. T. Unwin, E. Sherrard-Smith, T. S. Churcher, and A. C. Ghani, Quantifying the direct and indirect protection provided by insecticide treated bed nets against malaria, Nature Communications 14, 676 (2023).

[22] T. H. Barker, J. C. Stone, S. Hasanoff, C. Price, A. Kabaghe, and Z. Munn, Effectiveness of dual active ingredient insecticide-treated nets in preventing malaria: A systematic review and meta-analysis, PLoS One 18, e0289469 (2023).

[23] J. F. Mosha, M. A. Kulkarni, E. Lukole, N. S. Matowo, C. Pitt, L. A. Messenger, E. Mallya, M. Jumanne, T. Aziz,R. Kaaya, et al., Effectiveness and cost-effectiveness against malaria of three types of dual-active-ingredient long-lasting insecticidal nets (LLINs) compared with pyrethroid-only LLINs in Tanzania: a four-arm, cluster-randomised trial, The Lancet 399, 1227 (2022).

[24] H. M. Koenker, J. O. Yukich, A. Mkindi, R. Mandike, N. Brown, A. Kilian, and C. Lengeler, Analysing and recommending options for maintaining universal coverage with long-lasting insecticidal nets: the case of Tanzania in 2011, Malaria Journal 12, 1 (2013).

[25] A. D. Flaxman, N. Fullman, M. W. Otten Jr, M. Menon, R. E. Cibulskis, M. Ng, C. J. Murray, and S. S. Lim, Rapid scaling up of insecticide-treated bed net coverage in Africa and its relationship with development assistance for health: a systematic synthesis of supply, distribution, and household survey data, PLoS Medicine 7, e1000328 (2010).

[26] N. Golding, Spatiotemporal modelling of the spread of insecticide resistance in Africa, https://github.com/goldingn/ir_cube (2022).

[27] T. B. Knox, E. O. Juma, E. O. Ochomo, H. Pates Jamet, L. Ndungo, P. Chege, N. M. Bayoh, R. N’Guessan, R. N. Christian, R. H. Hunt, et al., An online tool for mapping insecticide resistance in major anopheles vectors of human malaria parasites and review of resistance status for the afrotropical region, Parasites & Vectors 7, 76 (2014).

[28] VAP, Vector Atlas (2026).

[29] WHO, Malaria Threat Mapper (2026).

[30] G. F. Killeen, F. O. Okumu, R. N’Guessan, M. Coosemans, A. Adeogun, S. Awolola, J. Etang, R. K. Dabiré, and V. Corbel, The importance of considering community-level effects when selecting insecticidal malaria vector products, Parasites & vectors 4, 160 (2011).

[31] R. K. Nash, B. Lambert, R. N’Guessan, C. Ngufor, M. Rowland, R. Oxborough, S. Moore, P. Tungu, E. Sherrard-Smith, and T. S. Churcher, Systematic review of the entomological impact of insecticide-treated nets evaluated using experimental hut trials in Africa, Current Research in Parasitology & Vector-borne Diseases 1, 100047 (2021).

[32] P. M. Gichuki, L. Kamau, K. Njagi, S. Karoki, N. Muigai, D. Matoke-Muhia, N. Bayoh, E. Mathenge, and R. S. Yadav, Bioefficacy and durability of Olyset® Plus a permethrin and piperonyl butoxide-treated insecticidal net in a 3-year long trial in Kenya, Infectious Diseases of Poverty 10, 16 (2021).

[33] K. R. Tan, J. Coleman, B. Smith, B. Hamainza, C. Katebe-Sakala, C. Kean, A. Kowal, J. Vanden Eng, T. K. Parris, C. T. Mapp, et al., A longitudinal study of the durability of long-lasting insecticidal nets in Zambia, Malaria Journal 15, 106 (2016).

[34] J. Raharinjatovo, R. K. Dabiré, K. Esch, D. D. Soma, A. Hien, T. Camara, M. B. Diouf, A. Belemvire, L. Gerberg, T. S. Awolola, et al., Physical and insecticidal durability of Interceptor®, Interceptor® G2, and PermaNet® 3.0 insecticide-treated nets in Burkina Faso: results of durability monitoring in three sites from 2019 to 2022, Malaria Journal 23, 173 (2024).

[35] J. L. Martin, L. A. Messenger, E. Bernard, M. Kisamo, P. Hape, O. Sizya, E. Festo, W. Matiku, V. Marcel, E. Malya, et al., Evaluation of bio-efficacy of field-aged novel long-lasting insecticidal nets (pbo, chlorfenapyr or pyriproxyfen combined with pyrethroid) against anopheles gambiae (ss) in Tanzania, Current Research in Parasitology & Vector-borne Diseases 6, 100216 (2024).

[36] J. L. Vanden Eng, A. Chan, A. P. Abílio, A. Wolkon, G. Ponce de Leon, J. Gimnig, and J. Morgan, Bed net durability assessments: exploring a composite measure of net damage, PLoS One 10, e0128499 (2015).

[37] A. Wheldrake, E. Guillemois, H. Arouni, V. Chetty, and S. J. Russell, The causes of holes and loss of physical integrity in long-lasting insecticidal nets, Malaria Journal 20, 45 (2021).

[38] J. T. Griffin, T. D. Hollingsworth, L. C. Okell, T. S. Churcher, M. White, W. Hinsley, T. Bousema, C. J. Drakeley,N. M. Ferguson, M.-G. Basáñez, et al., Reducing plasmodium falciparum malaria transmission in Africa: a model-based evaluation of intervention strategies, PLoS Medicine 7, e1000324 (2010).

[39] J. T. Griffin, S. Bhatt, M. E. Sinka, P. W. Gething, M. Lynch, E. Patouillard, E. Shutes, R. D. Newman, P. Alonso, R. E. Cibulskis, et al., Potential for reduction of burden and local elimination of malaria by reducing Plasmodium falciparum malaria transmission: a mathematical modelling study, The Lancet Infectious Diseases 16, 465 (2016).

[40] P. A. Eckhoff, A malaria transmission-directed model of mosquito life cycle and ecology, Malaria Journal 10, 303 (2011).

[41] T. Spangenberg, J. N. Burrows, P. Kowalczyk, S. McDonald, T. N. Wells, and P. Willis, The open access malaria box: a drug discovery catalyst for neglected diseases, PloS One 8, e62906 (2013).

[42] N. Chitnis, D. Hardy, and T. Smith, A periodically-forced mathematical model for the seasonal dynamics of malaria in mosquitoes, Bulletin of Mathematical Biology 74, 1098 (2012).

[43] W. A. Woldegerima, R. Ouifki, and J. Banasiak, Mathematical analysis of the impact of transmission-blocking drugs on the population dynamics of malaria, Applied Mathematics and Computation 400, 126005 (2021).

[44] C. Sangbakembi-Ngounou, C. Costantini, N. M. Longo-Pendy, C. Ngoagouni, O. Akone-Ella, N. Rahola, S. Cornelie, P. Kengne, E. R. Nakouné, N. P. Komas, et al., Diurnal biting of malaria mosquitoes in the Central African Republic indicates residual transmission may be “out of control”, Proceedings of the National Academy of Sciences 119, e2104282119 (2022).

[45] I. J. Stopard, T. S. Churcher, and B. Lambert, Estimating the extrinsic incubation period of malaria using a mechanistic model of sporogony, PLoS Computational Biology 17, e1008658 (2021).

[46] P. Brasil, A. de Pina Costa, R. S. Pedro, C. da Silveira Bressan, S. da Silva, P. L. Tauil, and C. T. Daniel-Ribeiro, Unexpectedly long incubation period of Plasmodium vivax malaria, in the absence of chemoprophylaxis, in patients diagnosed outside the transmission area in Brazil, Malaria Journal 10, 122 (2011).

[47] L. M. Lorenz, J. Bradley, J. Yukich, D. J. Massue, Z. Mageni Mboma, O. Pigeon, J. Moore, A. Kilian, J. Lines, W. Kisinza, et al., Comparative functional survival and equivalent annual cost of 3 long-lasting insecticidal net (LLIN) products in Tanzania: a randomised trial with 3-year follow up, PLoS Medicine 17, e1003248 (2020).

