## Supplementary Information for "Efficacy-adjusted use: modelling a refined metric of insecticide treated net coverage across Africa"

### Supplementary Information
### Efficacy adjusted insecticide treated nets coverage estimates from entomological inoculation rate surrogates

#### 1 Bioassay efficacy models

The efficacy of insecticidal agents in ITNs is commonly assessed using cone bioassay tests. These tests quantify efficacy based on mortality following ITN exposure within a given time period, with either mortality and knockdown rates being the most commonly reported. These measures give an approximate measure of ITN efficacy are representative of performance in ideal lab conditions, and do not actively track the waning of efficacy due to physical or chemical degradation.

We choose to model the waning bioefficacy of ITNs as a sigmoidal curve given by a Weibull model. To maintain functional independence from insecticide resistance effects, we assume the case where $\gamma = 1$ and thus model bioassay mortality as

$$\nu(t) = \nu_0 e^{-\left(\frac{t}{b}\right)^k}, \tag{1}$$

where $b$ and $k$ are model parameters to be fitted. Parameters are fit using a Bayesian approach with a mildly informative Gamma prior,

$$b \sim \text{Gamma}(9/2, 3/2), \tag{2}$$
$$k \sim \text{Gamma}(9/2, 3/2), \tag{3}$$

corresponding to a waning curve where there maximum rate of efficacy reduction occurs at approximately 2.6 years.

For calibration, we compiled data from four longitudinal studies of ITN bioefficacy [1–4]. These studies collectively track 24 hour cone bioassay mortality of a variety of LLIN, PBO and DAIs across 3 years in 4 different countries (Kenya, Zambia, Burkina Faso, Tanzania). The total compiled dataset consists of 46 (*time*,*efficacy*) observation pairs across 8 net brands representing conventional LLINs, and next-generation PBO and DAI nets. We choose to use reported values of 24 hour mortality rates as proxies for efficacy and assume Gaussian observational errors. Compiled data is provided in the Supplementary Material.

Fits of the efficacy decay curve $\nu(t)$ are given in Figure 1 with posterior estimate for model parameters of $b = 2.45$ (95 % marginal CrI = $[2.05, 2.75]$) and $k = 3.8$ (95 % marginal CrI = $[2.55, 9.9]$). Credible intervals are approximated by assuming a unimodal joint likelihood distribution and identifying the boundary of the sublevel set $S$ such that $\iint_S \mathcal{L}(b,k)\, db\, dk = 0.95$.

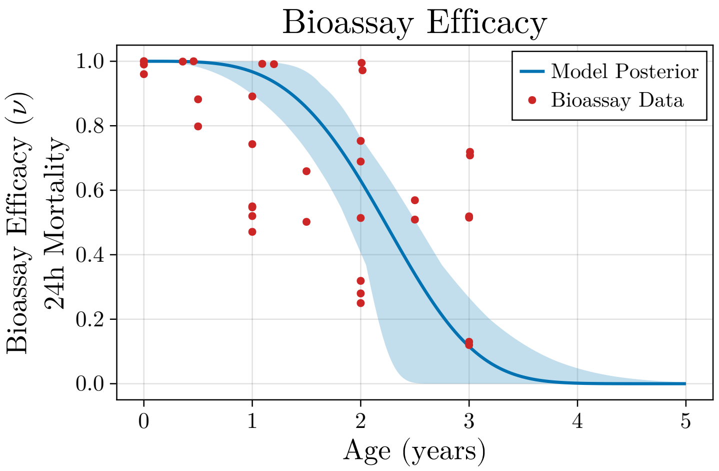


Figure 1: Posterior model fit of waning bioefficacy curve $\nu(t)$ with 95% credible interval shaded.

#### 2 EIR model solutions

Theoretical values of EIR in our analyses are calculated using the steady state solutions of a compartmental model of malaria in mosquito-human populations. The first model is evaluated in the absence of interventions and is given as follows with a simplified set of notation $\{k_i\}$ for the constant model parameters,

$$\frac{dS_m}{dt} = R(t) - k_3 S_m(t) I_h(t) - k_1 S_m(t), \tag{4a}$$

$$\frac{dL_m}{dt} = k_3 S_m(t) I_h(t) - k_4 L_m(t-\tau_m) L_m(t) - k_1 L_m(t), \tag{4b}$$

$$\frac{dI_m}{dt} = k_4 L_m(t-\tau_m) L_m(t) - k_2 I_m(t), \tag{4c}$$

$$\frac{dS_h}{dt} = k_5 I_h(t) - k_6 S_h(t) I_m(t), \tag{4d}$$

$$\frac{dL_h}{dt} = k_6 S_h(t) I_m(t) - L_h(t-\tau_h) L_h(t), \tag{4e}$$

$$\frac{dI_h}{dt} = L_h(t-\tau_h) L_h(t) - k_5 I_h(t). \tag{4f}$$

The steady-solution for the EIR model with no interventions is used to calculate a reference value for baseline mosquito reproduction rate at equilibrium. To do this, we assume the following population conditions:

- Mosquito and human populations are stationary: $\frac{d}{dt}(S_m + L_m + I_m) = \frac{d}{dt}(S_h + L_h + I_h) = 0$
- No substantial reductions in overall human populations due to death: $N_h = S_h + L_h + I_h$
- Mosquito populations settle at predetermined constant density: $N_{m,0} = S_m^\infty + L_m^\infty + I_m^\infty$.

For clarity of notation, $*$ refers to the steady-state values. Applying the above conditions yields the following reduced set of equations,

$$L_h^* = \sqrt{k_5 I_h^*}, \tag{5a}$$

$$S_h^* = 1 - L_h^* - I_h^*, \tag{5b}$$

$$I_m^* = \frac{L_h^{*2}}{k_6 S_h^*}, \tag{5c}$$

$$L_m^* = \sqrt{\frac{k_2 I_m^*}{k_4}}, \tag{5d}$$

$$S_m^* = \frac{k_1 L_m^* + k_2 I_m^*}{k_3 I_h^*}, \tag{5e}$$

$$N_{m,0} = S_m^* + L_m^* + I_m^*, \tag{5f}$$

subject to the conditions that all variables $\{S_m^*, L_m^*, I_m^*, S_h^*, L_h^*, SI_h^*\} \geq 0$. These equations can be numerically solved by optimising the following loss with respect to the variable $I_h^*$ using any choice of conventional optimisation algorithms (e.g. exhaustive search, golden search, bisection) to solve:

$$\min_{I_h^*} \| N_{m,0} - (S_m^* + L_m^* + I_m^*) \|. \tag{6}$$

Once calculated, the steady-state reproduction rate of mosquitoes can be calculated as,

$$R_{ss} = k_1(S_m^* + L_m^*) + k_1 I_m^*. \tag{7}$$

Solutions for the steady-state EIR model with interventions are derived using the same method.

#### 3 Steady state approximations for EIT model with interventions

To test the validity of the steady-state approximation of the EIR model, we construct a synthetic input time series for use $\xi(t)$ with a constant set mean net age. From this, two set of trajectories are calculated for the density infected of mosquitoes, which is a proxy for the theoretical EIR: $I_m(t)$ – density of infected mosquitoes based on the integration of the EIR model proper with respect to $\xi(t)$, and $I_m^\infty(t|\xi(t))$ – a time series of the steady state infected mosquito density.

For the synthetic dataset, we assume ITN mass campaign distributions at 5 year intervals and a net attrition curve following compact sigmoidal loss function of the MITN model with attrition parameters ($\tau = 10.85, \kappa = 19.44$) taken from the LLIN attrition parameters for Kenya (see [5]). For simplicity, the mean net age is assumed to be static at 1.5 years, and used to calculate $\eta_k, \eta_d$. Simulation results are given in Figure 2 and show that following a short transient period, the the steady state approximation infected mosquito density closely follows that of the full EIR solution.

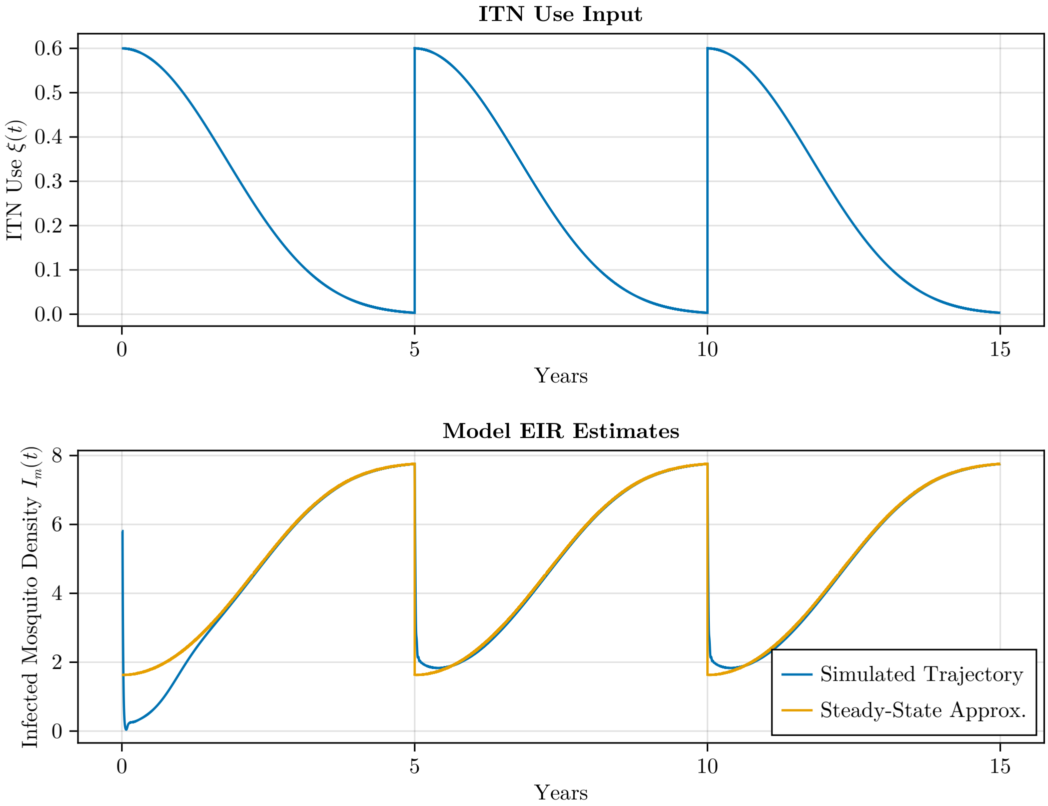


Figure 2: Trajectories of infected mosquito density for the full integrated EIR model, and the steady state approximation.

### 4 Barrier effect ($\eta_b$) calibration

Barrier effects of ITNs are represented by a single parameter $\eta_b$ which represents the probability of any given bite attempt from mosquito being mitigated due to the presence of a physical barrier (i.e. the bednet). Due to the difficulty in estimating true values for $\eta_b$, we instead calibrate the parameter $\eta_b$ based on reported model results by Unwin et al. [6]. These studies combine modelling outputs from the *malariasimulation* model [7] together with collected observational data to provide estimates of EIR reduction for various scenarios of use level $\xi$ and attempts to decompose ITN protection into barrier and insecticidal effects.

For calibration, we extract the three reported data points of use $\xi$ and resulting EIR reduction for the case where only barrier effects are present without insecticides. These point values are $\xi = [0.1, 0.5, 0.8]$ and $(1 - \frac{EIR(\xi)}{EIR_0}) = [0.05, 0.23, 0.36]$. Simple optimisation yields an optimal estimate of $\eta_b = 0.327$.

Two different validation tests are performed. For the first, we calculate the proposed model's ability to reproduce Unwin et al's estimates of EIR reduction at various use levels based purely on the Imperial model. Validation datapoints are $\xi = [0.1, 0.5, 0.8]$ and $(1 - \frac{EIR(\xi)}{EIR_0}) = [0.17, 0.66, 0.87]$, with our fitted value of $\eta_b = 0.327$ yielded a MSE of 0.068. For the second validation, we attempt to use our reduced ODE model to reconstruct the predicted $\xi$-EIR reduction curves by Unwin et al. for $\xi \in [0, 0.9]$ and find that the model closely reproduces the mean predictions as shown in Figure 7 of the main text.

### References


[1] P. M. Gichuki, L. Kamau, K. Njagi, S. Karoki, N. Muigai, D. Matoke-Muhia, N. Bayoh, E. Mathenge, and R. S. Yadav, "Bioefficacy and durability of Olyset® Plus, a permethrin and piperonyl butoxide-treated insecticidal net in a 3-year long trial in Kenya", Infectious Diseases of Poverty **10**, 16–26 (2021).

[2] K. R. Tan, J. Coleman, B. Smith, B. Hamainza, C. Katebe-Sakala, C. Kean, A. Kowal, J. Vanden Eng, T. K. Parris, C. T. Mapp, et al., "A longitudinal study of the durability of long-lasting insecticidal nets in Zambia", Malaria Journal **15**, 106 (2016).

[3] J. Raharinjatovo, R. K. Dabiré, K. Esch, D. D. Soma, A. Hien, T. Camara, M. B. Diouf, A. Belemvire, L. Gerberg, T. S. Awolola, et al., "Physical and insecticidal durability of Interceptor®, Interceptor® G2, and PermaNet® 3.0 insecticide-treated nets in Burkina Faso: results of durability monitoring in three sites from 2019 to 2022", Malaria Journal **23**, 173 (2024).

[4] J. L. Martin, L. A. Messenger, E. Bernard, M. Kisamo, P. Hape, O. Sizya, E. Festo, W. Matiku, V. Marcel, E. Malya, et al., "Evaluation of bio-efficacy of field-aged novel long-lasting insecticidal nets (pbo, chlorfenapyr or pyriproxyfen combined with pyrethroid) against anopheles gambiae (ss) in Tanzania", Current Research in Parasitology & Vector-borne Diseases **6**, 100216 (2024).

[5] E. Tan, M. van den Berg, C. Vargas, P. W. Gething, and T. L. Symons, "Dynamical and time series approach to understanding compartmental stock and flow models: a case study in malaria intervention models", Journal of the Royal Society Interface (2026).

[6] H. J. T. Unwin, E. Sherrard-Smith, T. S. Churcher, and A. C. Ghani, "Quantifying the direct and indirect protection provided by insecticide treated bed nets against malaria", Nature Communications **14**, 676 (2023).

[7] J. T. Griffin, T. D. Hollingsworth, L. C. Okell, T. S. Churcher, M. White, W. Hinsley, T. Bousema, C. J. Drakeley, N. M. Ferguson, M.-G. Basáñez, et al., "Reducing plasmodium falciparum malaria transmission in Africa: a model-based evaluation of intervention strategies", PLoS Medicine **7**, e1000324 (2010).
